# Artemether-lumefantrine-amodiaquine or artesunate-amodiaquine combined with single low-dose primaquine to reduce *Plasmodium falciparum* malaria transmission in Ouélessébougou, Mali: a five-arm, phase 2, single-blind, randomised clinical trial

**DOI:** 10.1101/2024.02.23.24303266

**Authors:** Almahamoudou Mahamar, Leen N Vanheer, Merel J Smit, Koualy Sanogo, Youssouf Sinaba, Sidi M. Niambele, Makonon Diallo, Oumar M Dicko, Richard S. Diarra, Seydina O Maguiraga, Ahamadou Youssouf, Adama Sacko, Sekouba Keita, Siaka Samake, Adama Dembele, Karina Teelen, Yahia Dicko, Sekou F. Traore, Arjen Dondorp, Chris Drakeley, William Stone, Alassane Dicko

**Affiliations:** Malaria Research and Training Centre, Faculty of Pharmacy and Faculty of Medicine and Dentistry, University of Sciences Techniques and Technologies of Bamako, Bamako, Mali; Department of Infection Biology, London School of Hygiene & Tropical Medicine, London, UK, WC1E7HT; Department of Medical Microbiology and Radboud Center for Infectious Diseases, Radboud University Medical Center, Nijmegen, the Netherlands; Mahidol-Oxford Tropical Medicine Research Unit, Faculty of Tropical Medicine, Mahidol University, Bangkok 10400, Thailand; Centre for Tropical Medicine and Global Health, Nuffield Department of Clinical Medicine, University of Oxford, Oxford, UK

**Keywords:** Gametocytes, Artemether-lumefantrine, Primaquine, Artemether-lumefantrine-Amodiaquine, Triple Artemisinin-based Combination Therapy, Artesunate-Amodiaquine, Transmission-blocking

## Abstract

**Background:** Triple artemisinin-based combination therapies, such as artemether-lumefantrine-amodiaquine, can delay the spread of antimalarial drug resistance; artesunate-amodiaquine is widely used for uncomplicated *Plasmodium falciparum* malaria. We aimed to determine the efficacy of artemether-lumefantrine-amodiaquine and artesunate-amodiaquine with and without single low-dose primaquine for reducing gametocyte carriage and transmission to mosquitoes.

**Methods:** We conducted a five-arm, single-blind, phase 2, randomised clinical trial at the Ouélessébougou Clinical Research Unit of the Malaria Research and Training Centre of the University of Sciences, Techniques and Technologies of Bamako (Bamako, Mali). Eligible participants aged 10-50 years, with asymptomatic *P. falciparum* microscopy-detected gametocyte carriage, were randomised (1:1:1:1:1) to receive either artemether-lumefantrine, artemether-lumefantrine-amodiaquine, artemether-lumefantrine-amodiaquine plus primaquine, artesunate-amodiaquine, or artesunate-amodiaquine plus primaquine. Treatment allocation was computer randomised and concealed to all study staff other than the trial pharmacist. The primary outcome was the within-person percentage reduction in mosquito infection rate at 48 hours after treatment initiation compared to pre-treatment, assessed by direct membrane feeding assay. Data were analysed per protocol. This study is registered with ClinicalTrials.gov, NCT05550909.

**Findings:** Between Oct 16 and Dec 28, 2022, 1249 individuals were screened for eligibility, 100 of which were enrolled and randomly assigned to one of five treatment groups (n=20 per group). Before treatment, 61 (61%) of 100 participants were infectious to mosquitoes, with a median of 7·3% (IQR 3·2-23·5) of mosquitoes becoming infected. Among infectious individuals, the median percentage reduction in mosquito infection rate between pre-treatment and 2 days post-treatment was 100% (IQR 100-100) in the artemether-lumefantrine (p=0·0018), artemether-lumefantrine-amodiaquine (p=0·0018), and artemether-lumefantrine-amodiaquine plus primaquine (p=0·0009) treatment groups. In the artesunate-amodiaquine group the median percent reduction in mosquito infection rate was only *31·67% (IQR -10·9-100, p=0*·1927), whereas there was 100% reduction in the artesunate-amodiaquine plus primaquine group (p=0·0009). At day 2, 10% (2/20) of participants in the artemether-lumefantrine group, 11% (2/19) in the artemether-lumefantrine-amodiaquine group, and 75% (15/20) in the artesunate-amodiaquine group infected any number of mosquitoes whilst no infected mosquitoes were observed at this time-point in the primaquine arms. No serious adverse events occurred.

**Interpretation:** These data support the effectiveness of artemether-lumefantrine alone or as part of triple combination therapy for preventing nearly all human-mosquito malaria parasite transmission within 48 hours. In contrast, substantial transmission was observed following treatment with artesunate-amodiaquine. The addition of a single low-dose of primaquine blocks transmission to mosquitoes rapidly regardless of schizonticide.

**Funding:** Bill & Melinda Gates Foundation

## INTRODUCTION

Malaria morbidity and mortality remains unacceptably high (1) and the emergence and spread of partial resistance against artemisinin derivatives, the main component of artemisinin-based combination therapies (ACTs), in South East Asia (2,3) and East-Africa (4,5) is threatening to increase malaria cases and deaths. There is therefore a clear need for antimalarial treatments designed to slow the spread of resistance, either through novel combinations of existing drugs or supplementation with drugs that have specific effects on gametocytes, the sexual life stages responsible for maintaining parasite transmission. For optimal use, it is essential that we understand how effectively current and future antimalarials combat gametocytes, and how this translates into reductions in transmission to mosquitoes.

Triple artemisinin-based combination therapies (TACT) combine an existing ACT with a second partner drug that is slowly eliminated, to reduce the likelihood of incomplete parasite clearance and thus delay the spread of artemisinin resistance (6). Artemether-lumefantrine-amodiaquine is a TACT that has been proven safe, well-tolerated and efficacious for the treatment of uncomplicated *P. falciparum* malaria, including in areas with artemisinin and partner drug resistance (7,8). The effect of artemether-lumefantrine-amodiaquine on mature gametocytes and infectivity is unknown. Artesunate-amodiaquine is the first-line ACT for uncomplicated *P. falciparum* malaria in many countries (9), but its transmission reducing efficacy has not been tested directly. Studies assessing gametocyte carriage after artesunate-amodiaquine observed persistent gametocyte carriage post-treatment for 21 days or longer, but without transmission assays the infectivity of these persisting gametocytes cannot be confirmed (10–12).

Although artemisinin-based treatments have superior gametocytocidal properties to non-artemisinin’s (13), with artemether-lumefantrine being the most potent (14), the transmission reducing activities of ACTs vary widely (14–16). In contrast, the 8-aminoquinoline primaquine is a potent gametocytocidal drug which at a single low-dose (0·25mg/kg) blocks transmission within 48 hours of treatment. The World Health Organization (WHO) recommends the addition of a single low-dose of primaquine (0·25 mg/kg) to ACT to reduce *P. falciparum* transmission. The gametocytocidal and transmission reducing activities of single low-dose primaquine have been assessed in combination with dihydroartemisinin-piperaquine, pyronaridine-artesunate and artemether-lumefantrine (14–18), however, combining artemether-lumefantrine-amodiaquine or artesunate-amodiaquine with a single low-dose primaquine for *P. falciparum* transmission reduction has not yet been tested.

In the current study, we aimed to determine the safety and efficacy of artemether-lumefantrine-amodiaquine and artesunate-amodiaquine with and without single low-dose primaquine for reducing the transmission of *P. falciparum* gametocytes in a cohort of Malian children and adults. In addition, to investigate the mechanisms of artemether-lumefantrine’s superior transmission reducing efficacy, we employed magnetic-activated cell sorting (MACS) to enrich the gametocyte content of the mosquito blood meal source in our transmission assays, allowing us to differentiate between lack of infectivity due to insufficient gametocyte densities and drug-induced sterilization effects.

## METHODS

### Study design and participants

This five-arm, single-blind, phase 2 randomised controlled trial was conducted at the Ouélessébougou Clinical Research Unit of the Malaria Research and Training Centre (MRTC) of the University of Sciences, Techniques and Technologies of Bamako in Mali. Ouélessébougou is a commune that includes the town of Ouélessébougou and 44 surrounding villages, which have a total of approximately 50,000 inhabitants. Malaria transmission is highly seasonal, tied to the rainy season occurring from July to November. The prevalence of *P. falciparum* malaria and gametocytes in children aged over 5 years varies between 50-60% and 20-25%, respectively, during the transmission season. The study team met with community leaders, village health workers, and heads of households from each village, before the commencement of screening, to explain the study and obtain approval. Village health workers then used a door-to-door approach to inform households of the date and location where consenting and screening would take place. Participants were included in the trial if they met the following criteria: positive for *P. falciparum* gametocytes by microscopy (i.e. ≥1 gametocytes observed in a thick film against 500 white blood cells (WBC), equating to 16 gametocytes/µL with a standard conversion of 8000 WBC)/µL blood); absence of other non-*P. falciparum* species on blood film; haemoglobin density of ≥10 g/dL; aged between 10-50 years; bodyweight of ≤80 kg; no clinical signs of malaria, defined by fever (≥37·5°C); no signs of acute, severe or chronic disease. Exclusion criteria included pregnancy (tested at enrolment by urine test) or lactation, allergies to any of the study drugs, use of other medication (except for paracetamol and/or aspirin), use of antimalarial drugs over the past week, history of prolongation of the corrected QT (QTc) interval, documented or self-reported history of cardiac conduction problems or epileptic seizures, and blood transfusion in last 90 days. Prior to screening and prior to study enrolment, participants provided written informed consent (≥18 years) or written parental consent (10-17 years). In addition to parental consent, an assent was sought for 12-17 years old. Ethical approval was granted by the Ethics Committee of the University of Sciences, Techniques, and Technologies of Bamako (Bamako, Mali) (No2022/244/CE/USTTB), and the Research Ethics Committee of the London School of Hygiene and Tropical Medicine (London, United Kingdom) (LSHTM Ethics Ref. 28014).

### Randomisation and masking

Allocation to five treatment groups (artemether-lumefantrine, artemether-lumefantrine-amodiaquine, artemether-lumefantrine-amodiaquine plus primaquine, artesunate-amodiaquine and artesunate-amodiaquine plus primaquine) was randomised in a 1:1:1:1:1 ratio. Enrolment continued until 100 participants were enrolled (20 individuals assigned to each treatment group). An independent MRTC statistician randomly generated the treatment assignment using Stata version 16 (StataCorp, College Station, Texas, USA), which was linked to participant ID number. The statistician prepared sealed, opaque envelopes with the participant ID number on the outside and treatment assignment inside which were sent to the MRTC study pharmacist. The study pharmacist provided treatment and was consequently not blinded to treatment assignment; staff involved in assessing safety, infectivity and laboratory outcomes were blinded.

### Procedures

Artesunate-amodiaquine and artemether-lumefantrine treatment (Guilin Pharmaceutical, Shanghai, China) was administered over three days as per manufacturer instructions (supplementary information 1). Participants in the artemether-lumefantrine plus amodiaquine group were treated with standard doses of artemether-lumefantrine and amodiaquine (Guilin Pharmaceutical, Shanghai, China) as per manufacturer instructions. A single dose of 0·25 mg/kg primaquine (ACE Pharmaceuticals, Zeewolde, The Netherlands) was administered on day 0 in parallel with the first dose of ACT or TACT, as described previously (18).

Participants received a full clinical and parasitological examination on days 2, 7, 14, 21, and 28 after receiving the first dose of the study drugs (supplementary figure 1). Giemsa-stained thick film microscopy was performed as described previously, with gametocytes counted against 500 WBC and asexual stages counted against 200 WBC 23/02/2024 08:44:00. Total nucleic acids were extracted for molecular gametocyte quantification using a MagNAPure LC automated extractor (Total Nucleic Acid Isolation Kit-High Performance; Roche Applied Science, Indianapolis, IN, USA). Female and male gametocytes were quantified in a multiplex reverse transcriptase quantitative PCR (RT-qPCR) assay (supplementary table 1) (19). Samples were classified as negative for a particular gametocyte sex if the qRT-PCR quantified density of gametocytes of that sex was less than 0·01 gametocytes per μL (i.e. one gametocyte per 100 μL of blood sample). Haemoglobin density was measured using a haemoglobin analyser (HemoCue; AB Leo Diagnostics, Helsingborg, Sweden) or automatic haematology analyser (HumaCount 5D; Wiesbaden, Germany). Additional venous blood samples were taken for biochemical and infectivity assessments on day 0, 2, 7, and 14 in all treatment groups. Levels of aspartate transaminase (AST), alanine transaminase (ALT) and blood creatine were measured using the automatic biochemistry analyser Human 100 (Wiesbaden, Germany). For each assessment of infectivity, ∼75 locally insectary-reared female *Anopheles gambiae* mosquitoes were allowed to feed for 15-20 minutes on venous blood samples (Lithium Heparin VACUETTE tube, Greiner Bio-One, Kremsmünster, Austria) through a prewarmed glass membrane feeder system (Coelen Glastechniek, Weldaad, the Netherlands). Mosquitoes that had taken no bloodmeal or a partial blood meal were discarded; surviving blood-fed mosquitoes were dissected on the 7th day post-feeding. Midguts were stained with 1% mercurochrome and examined for the presence and density of oocysts by expert microscopists.

Gametocytes in the infected whole blood sample were concentrated by MACS using a QuadroMACS™ separator and LS MACS columns (MiltenyiBiotech, UK) as previously described (20). Briefly, MACS LS columns were equilibrated with 1 mL of warm incomplete medium, followed by 3 mL of infected whole blood and 2 mL medium wash. LS columns were then removed from the magnet, and gametocytes were eluted in 4 mL of warm medium. Flow-through and gametocyte fractions were then centrifuged (2000 RPM, 5 minutes, 37°C). Medium was removed carefully, and the gametocyte pellet was resuspended in 450 µL warm malaria naïve serum and 600 µL of the same participants packed cells. The entire MACS procedure was carried out in a 37°C cabinet incubator.

### Outcomes

The primary outcome measure was median percentage change in mosquito infection rate between pre-treatment and 2 days after treatment initiation. Secondary outcomes were mosquito infection metrics (infectious individuals, mosquito infection rate and oocyst density) at prespecified timepoints (days 0, 2, 7 and 14); gametocyte and asexual parasite prevalence, density, gametocyte circulation time, area under the curve (AUC) of gametocyte density over time, and sex ratio (i.e., proportion of gametocytes that were male or female); and safety assessments including incidence of clinical and laboratory adverse events. Differences in all transmission metrics, gametocyte, asexual stages, and safety outcomes were compared between matched treatment groups (ie, artemether–lumefantrine vs artemether–lumefantrine-amodiaquine and artemether–lumefantrine-amodiaquine plus primaquine, artesunate-amodiaquine vs artesunate-amodiaquine plus primaquine) as secondary outcomes. Primary and secondary analyses of mosquito infection rate and oocyst density metrics were performed on individuals infectious at baseline, but are available for all individuals in the appendix. Exploratory outcomes included mosquito infection metrics after gametocyte enrichment, for within and between treatment group comparison. Gametocyte infectivity was assessed as an exploratory outcome using logistic regression models adjusted for gametocyte density, wherein the shape of the relationship between gametocyte density and mosquito infection rate was estimated using fractional polynomials.

Adverse events were graded by the study clinician for severity (mild, moderate, or severe) and relatedness to study medication (unrelated or unlikely, possibly, probably, or definitely related). A reduction in haemoglobin concentration of 40% or more from baseline was categorised as a haematological severe adverse event. An external data safety and monitoring committee was assembled before the trial. Safety data were discussed after enrolment of 50 participants, and after the final follow-up visit of the last participant.

### Statistical analysis

Sample size was informed by previous trials in the study setting using a mixed effects logistic regression model that accounted for correlation between mosquito observations from the same participant (15–18) and expecting a reduction in infectivity of 90% as previously detected for a single low-dose of primaquine (15,18). When including 20 participants per group and dissecting 50 mosquitoes per participant per timepoint, we calculated 92% empirical power to detect >85% reduction in infectivity with a one-tailed test with an α=0·05 level of significance. The sample size was not designed for between-group comparisons and any comparison of transmission-blocking effects between groups is secondary and limited to matched treatment groups. Mosquito infectivity was assessed at three levels: the percentage of participants infectious to any number of mosquitoes (ie, infectious individuals), the proportion of mosquitoes infected with any number of oocysts (ie, mosquito infection rate), and the mean number of oocysts in a sample of mosquitoes (ie, oocyst density).

Mosquito infection rate and oocyst density were analysed at timepoints after baseline only for those individuals who were infectious at baseline. The proportion of infectious individuals and the prevalence of gametocytes and asexual stage parasites were compared within and between treatment groups using one-sided Fishers exact tests. Mosquito infection rate was compared within-groups (relative to baseline) by Wilcoxon sign rank test (z-score) and between-groups by linear regression adjusted for baseline mosquito infection rate (t score, coefficient with 95% CI). For all direct membrane feeding assays (prior and post-gametocyte enrichment), the proportion of infectious individuals was compared between-group (direct membrane feeds prior to gametocyte concentration as reference) and within-group (relative to baseline) using one-sided Fishers exact tests. Haemoglobin levels were compared using paired t tests (t score) for within-group analyses and linear regression adjusted for baseline levels of each measure for between-group analyses (t score, coefficient with 95% CI). Percentage change from baseline was analysed using two-sample t tests for between group analysis and paired t tests (t score) for within-group analysis.

The proportion of gametocytes that were male was analysed for all values with total gametocyte densities of 0·2 gametocytes per μL or more, ensuring accurate quantification of sex ratios. Gametocyte circulation time was calculated to determine the mean number of days that a mature gametocyte circulates in the blood before clearance, using a deterministic compartmental model that assumes a constant rate of clearance and has a random effect to account for repeated measures on individuals, as described previously (21); difference in circulation time between groups and between gametocyte sexes was analysed using t tests (t score). Area under the curve (AUC) of gametocyte density per participant over time was calculated using the linear trapezoid method and was analysed by fitting linear regression models to the log10 adjusted AUC values, with adjustment for baseline gametocyte density (t score, coefficient with 95% CI). All other analyses of quantitative data were done using Wilcoxon sign rank tests (z-score) and Wilcoxon rank-sum tests (z-score). All comparisons were defined before study completion and analyses were not adjusted for multiple comparisons. For all analyses, the threshold for statistical significance was set at p<0·05.

Statistical analysis was conducted using Stata (version 17.0) and SAS (version 9.4). Data visualisation was performed using the R-based *ggplot2* package (R version 4.3.2) and Stata-based graphics (version 17.0). The trial is registered with ClinicalTrials.gov, NCT05550909.

## RESULTS

Between Oct 16 and Dec 28, 2022, 1249 individuals were screened for eligibility, 100 of whom were enrolled and randomly assigned to one of five treatment groups (n=20 per group; figure 1). Participant characteristics were similar between the study groups, although the proportion of infectious participants at baseline was higher in the artesunate-amodiaquine group (table 1). The primary outcome was recorded on day 2 of follow-up, with 98 (98%) of 100 individuals completing this study visit (one in the artemether-lumefantrine-amodiaquine group and one in the artemether-lumefantrine-amodiaquine plus primaquine group did not complete this visit). 96 (96%) participants completed all visits to day 28 (two in the artemether-lumefantrine-amodiaquine, one in the artemether-lumefantrine-amodiaquine plus primaquine, and one in the artesunate-amodiaquine plus primaquine group did not complete all visits).

**Figure 1.**
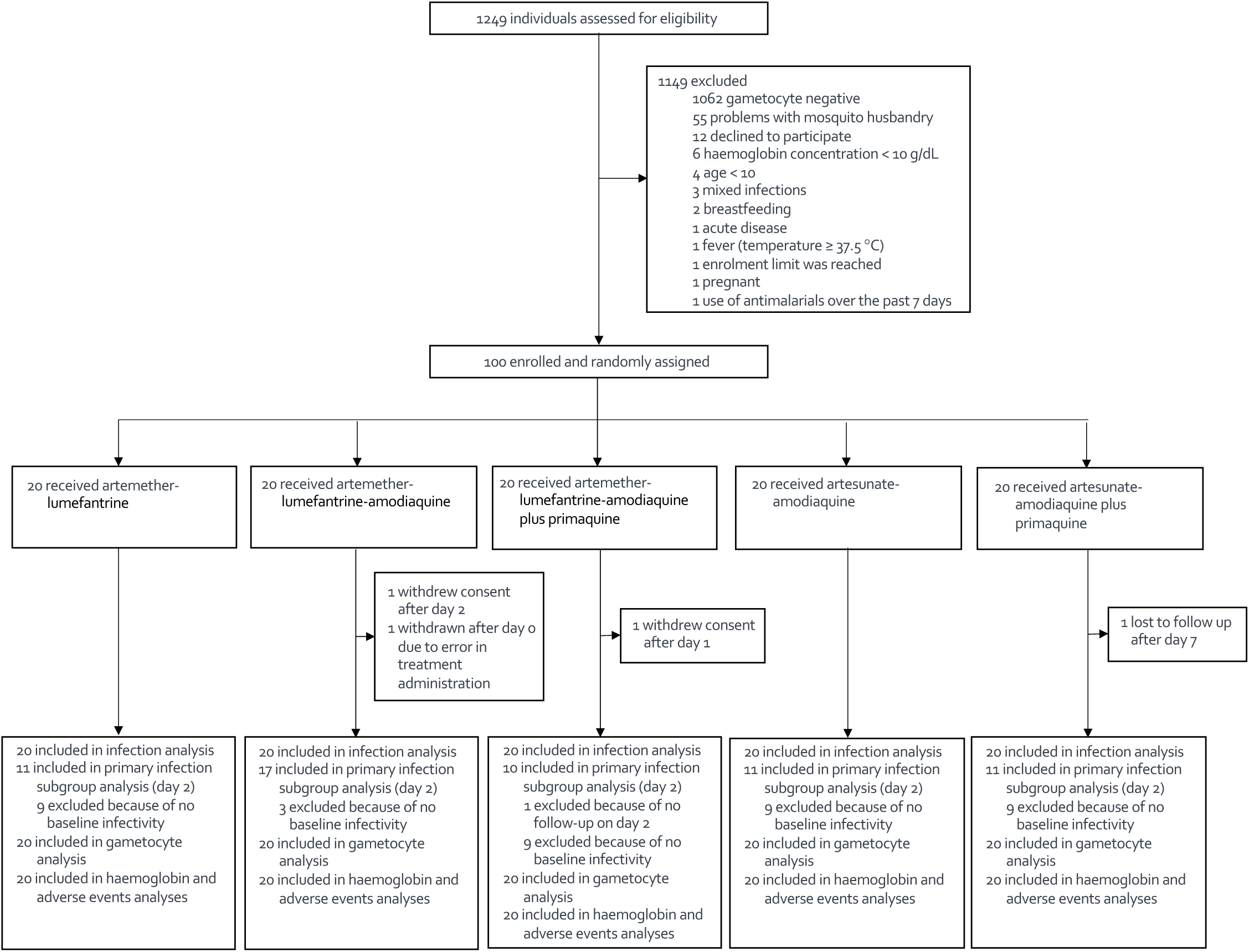
Trial profile. 96 of 100 (96%) participants completed all visits to day 28 (two in the artemether-lumefantrine-amodiaquine, one in the artemether-lumefantrine-amodiaquine plus primaquine, and one in the artesunate-amodiaquine plus primaquine group did not complete all visits).

**Table 1.** Characteristics of the study participants prior to treatment. Age, haemoglobin concentration and parasite densities are given as median and interquartile range [IQR]. Gametocyte prevalence and density were calculated from reverse transcriptase quantitative PCR targeting CCP4/PfMGET mRNA (gametocytes). Asexual parasite prevalence and density were assessed by thick film microscopy.

|  | AL (n=20) | AL-AQ (n=20) | AL-AQ+PQ (n=20) | AS-AQ (n=20) | AS-AQ+PQ (n=20) |
| --- | --- | --- | --- | --- | --- |
| Age in years (median [IQR]) | 13·0 (11·0-18·5) | 13·0 (11·5-28·0) | 13·0 (11·0-15·0) | 12·0 (10·0-16·0) | 12·5 (11·5-20·0) |
| Female participants | 11 (55%) | 10 (50%) | 12 (60%) | 11 (55%) | 7 (35%) |
| Male participants | 9 (45%) | 10 (50%) | 8 (40%) | 9 (45%) | 13 (65%) |
| Haemoglobin (g/dL) | 11·8 (11·5-13·8) | 12·1 (11·3-12·6) | 11·8 (11·1-12·1) | 11·4 (10·9-12·3) | 11·7 (11·2-12·8) |
| Gametocyte prevalence (n/% positive) | 20 (100%) | 20 (100%) | 20 (100%) | 20 (100%) | 20 (100%) |
| Gametocyte density (parasites/μL) | 31·0 (19·9-92·7) | 28·6 (11·5-130·5) | 42·3 (11·8-97·0) | 52·5 (33·6-129·0) | 24·8 (10·7-115·2) |
| Asexual parasite prevalence (n/% positive) | 10 (50%) | 6 (30%) | 5 (25%) | 8 (40%) | 8 (40%) |
| Asexual parasite density (parasites/μL) | 37·9 (0·0-300·0) | 0·0 (0·0-79·8) | 0·0 (0·0-37·6) | 0·0 (0·0-1654·9) | 0·0 (0·0-720·0) |

The median number of mosquitoes dissected in an individual mosquito feeding experiment was 60 (IQR 54-64). Before treatment, 61 (61%) individuals were infectious to mosquitoes, with a median of 7·3% (IQR 3·2–23·5) of mosquitoes becoming infected. The median number of oocysts per infected mosquito was 1·3 (IQR 1-3). At day 2 there was a significant within-person reduction in mosquito infection rate relative to baseline in all groups except for the artesunate-amodiaquine group. No individuals remained infectious to mosquitoes at day 2 in the treatment groups with primaquine (table 2). At the same timepoint, 2 (10%) of 20 participants in the artemether-lumefantrine group, 2 (11%) of 19 in the artemether-lumefantrine-amodiaquine group, and 15 (75%) of 20 in the artesunate-amodiaquine group infected any number of mosquitoes. At all timepoints after day 2, infectious participants were only found in the artesunate-amodiaquine group; 7 (35%) of 20 at day 7, 3 (16%) of 19 at day 14 and 1 (5%) of 19 at day 28 (figure 2 and supplementary table 2). Mosquito infection data for all individuals, regardless of baseline infectivity, is presented in supplementary table 3.

**Figure 2.**
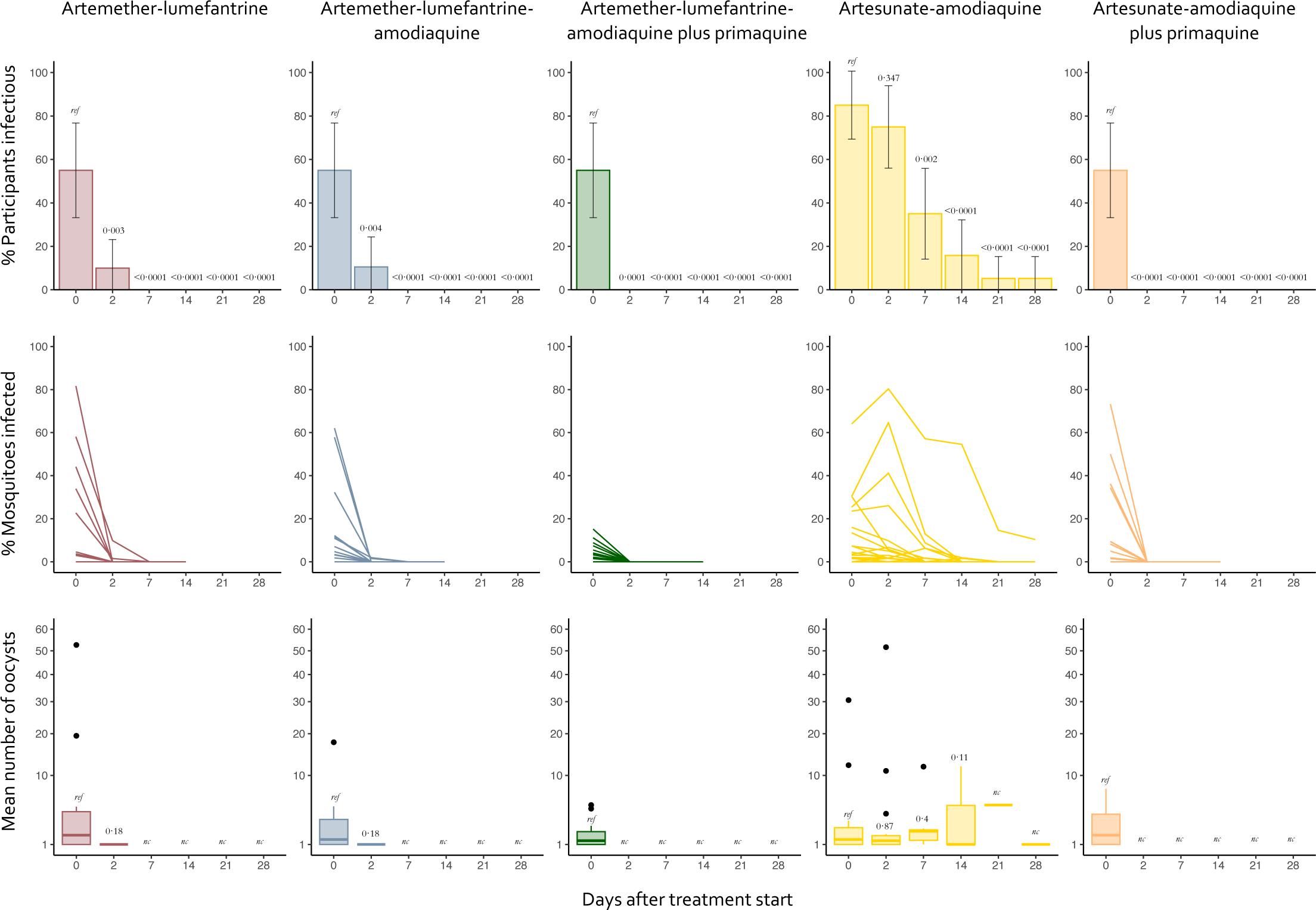
Participant infectivity and proportion of mosquitoes infected in direct membrane feeding assays. Error bars show 95% CI. The denominator for participants infectious is the total number of participants still enrolled at the given timepoint, rather than the number tested for infectivity at that timepoint. Infectivity assays were discontinued after 14 days when a participant did not infect any mosquitoes at two subsequent timepoints and were thereafter considered non-infectious. Each line in the plots showing percentage of mosquitoes infected represents an individual. Timepoints not indicated on the x-axis were not tested. The prevalence infectious individuals were compared within treatment groups using one-sided Fishers exact tests, and the mean number of oocysts were compared to baseline within-group by Wilcoxon rank-sum test.

**Table 2.**
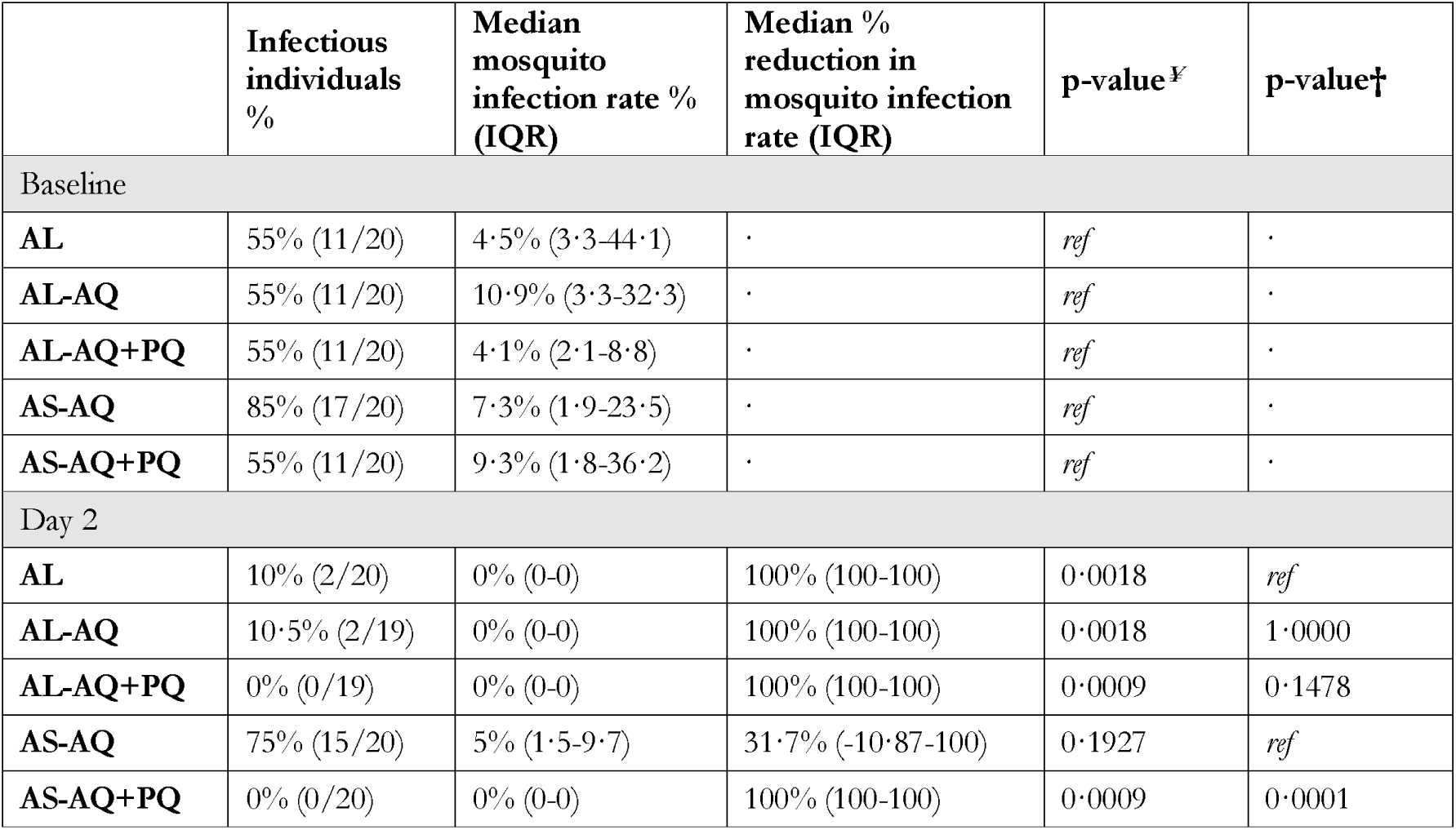
Median percent reduction in mosquito infection rate for individuals infectious before treatment. Individuals were classed as infectious if direct membrane feeding assays resulted in at least one mosquito with any number of oocysts. All values are for individuals who were infectious to mosquitoes before treatment. ¥ Within-group comparison of median reduction in mosquito infection rate by Wilcoxon signed rank test (day 0 as reference, primary outcome). †Between-group comparison of median reduction in mosquito infection rate (i.e., artemether–lumefantrine vs artemether–lumefantrine-amodiaquine and artemether–lumefantrine-amodiaquine plus primaquine, artesunate-amodiaquine vs artesunate-amodiaquine plus primaquine) by Wilcoxon rank-sum test. Full details of mosquito feeding assay outcomes are in supplementary table 2.

Gametocyte enrichment by MACS was performed on 95 blood samples collected pre-treatment and 94 blood samples that were collected on the second day after treatment initiation. Overall, gametocyte enrichment increased mosquito infection rates by a mean of 7·29% (supplementary figure 2). Comparing direct membrane feeds prior to enrichment with those after, the percentage of infectious individuals increased in the enrichment-boosted group in all treatment groups (supplementary figure 2, supplementary table 4), whilst at day 2, the percentage of infectious individuals increased for all treatment groups except for artemether-lumefantrine-amodiaquine plus primaquine, in which all individuals remained non-infectious. In the artesunate-amodiaquine plus primaquine group, two initially non-infectious individuals infected 1-4 mosquitoes following gametocyte enrichment.

Asexual parasite densities, measured by microscopy, decreased rapidly after treatment initiation, with only one individual in both the artemether-lumefantrine and artesunate-amodiaquine groups retaining asexual stages at day 2, whereas in all other treatment groups, no asexual parasites were observed after treatment initiation (supplementary table 5). Gametocyte densities declined over time in all treatment groups, though much more rapidly in those who received primaquine (supplementary figure 3, supplementary table 6). 18 (95%) of 20 participants treated with artesunate-amodiaquine were still gametocyte positive (> one gametocyte per 100 μL) at the final day of follow-up (day 28), whereas 16 (80%) of 20 in the artemether-lumefantrine and 13 (72%) of 18 in the artemether-lumefantrine-amodiaquine groups remained gametocyte positive at the same timepoint. Only one individual in both primaquine treatment groups had persisting gametocytes at day 28 (1 (5%) of 18 in the artemether-lumefantrine-amodiaquine plus primaquine and 1(5%) of 20 in the artesunate-amodiaquine plus primaquine groups). Total gametocyte circulation time was estimated at 6·1 days (95% CI 5·4-6·9) in the artemether-lumefantrine group, 6·0 days (5·2-6·8) in the artemether-lumefantrine-amodiaquine group and 2·6 (2·1-3·1) in the artemether-lumefantrine-amodiaquine plus primaquine group (supplementary table 7); the same measure was estimated at 7·9 days (6·7-9·3) and 3·3 days (2·8-3·8) in the artesunate-amodiaquine and artesunate-amodiaquine plus primaquine respectively. Gametocyte sex ratios showed a male bias from day 2 after treatment start in the artemether-lumefantrine, artemether-lumefantrine-amodiaquine and artemether-lumefantrine-amodiaquine plus primaquine groups and from day 7 in the artesunate-amodiaquine plus primaquine group, with significantly more males in the artesunate-amodiaquine plus primaquine group compared to the artesunate-amodiaquine alone group (supplementary tables 6 and 8, supplementary figure 4). Too few gametocytes persisted to make conclusions about absolute per-gametocyte infectivity (supplementary table 9).

There was a statistically significant within-group reduction in mean haemoglobin in all treatment groups at day 2 compared to baseline, however, by day 7 the haemoglobin levels had normalised in all group and were comparable to baseline (supplementary table 10, supplementary figure 5). The greatest reduction in mean haemoglobin density in any treatment group or timepoint was 5·58% (95% CI 3·64 to 7·54) in the artesunate-amodiaquine group at day 2. The were no statistically significant decreases in percent change in haemoglobin compared to baseline between treatment groups at any timepoint. The greatest reduction in haemoglobin density in any individual was 25·2% (from 14·3 g/dL at baseline to 10·7 g/dL at day 21 in an individual in the artemether-lumefantrine group). The lowest observed haemoglobin density in any individual and timepoint was 9 g/dL at baseline in an individual in the artesunate-amodiaquine group. No severe laboratory abnormalities occurred; all possibly drug related laboratory abnormalities normalised on the subsequent visit (supplementary table 11).

Overall, 85 (85%) of 100 participants had a total of 262 adverse events during follow-up, of which 181 (69%) were categorised as mild and 81 (30·1%) as moderate (table 3, supplementary table 12). No severe adverse events or serious adverse events occurred during the trial. The most common treatment-related adverse event was mild or moderate headache, which occurred in 43 (43%) participants (artemether-lumefantrine n=6; artemether-lumefantrine plus amodiaquine n=9; artemether-lumefantrine plus amodiaquine plus primaquine n=11; artesunate-amodiaquine n=8; and artesunate-amodiaquine plus primaquine n=9). There were no significant differences between treatment groups in the proportion of participants who experienced any mild adverse event (p=0·612), mild (p=0·178) or moderate (p=0·055) treatment-related adverse events at any study visit.

**Table 3.** Frequency of adverse events. Number of participants per group who experienced adverse events. Classification as ‘related to treatment’ was defined as probably, possibly or definitely related to treatment, as described in the methods. If there were multiple episodes per participant, the highest grade and most likely related to treatment is presented in this table. P-values are from Fisher’s exact tests for differences in proportion of individuals with an AE between all groups* or between AL-AQ or AL-AQ+PQ groups and the AL reference group and AS-AQ+PQ group and the AS-AQ reference group**. ref = reference group, nc = not calculable. AL = artemether-lumefantrine; AL-AQ = artemether-lumefantrine-amodiaquine; AL-AQ+PQ = artemether-lumefantrine-amodiaquine plus primaquine; AS-AQ = artesunate-amodiaquine; AS-AQ+PQ = artesunate-amodiaquine plus primaquine

| Description | Total (n=100) | AL (n=20) | AL-AQ (n=20) | AL-AQ+PQ (n=20) | AS-AQ (n=20) | AS-AQ+PQ (n=20) |
| --- | --- | --- | --- | --- | --- | --- |
| All | 85% (85/100) | 90% (18/20) | 75% (15/20) | 90% (18/20) | 80% (16/20) | 90% (18/20) |
| P-value | 0·612* | *ref* | 0·407** | 1·000** | *ref* | 0·661** |
| Mild related AE | 47% (47/100) | 60% (12/20) | 35% (7/20) | 40% (8/20) | 35% (7/20) | 65% (13/20) |
| P-value | 0·178* | *ref* | 0·205** | 0·343** | *ref* | 0·113** |
| Moderate related AE | 26% (26/100) | 10% (2/20) | 35% (7/20) | 35% (7/20) | 40% (8/20) | 10% (2/20) |
| P-value | 0·055* | *ref* | 0·127** | 0·127** | *ref* | 0·065** |
| Severe related AE | 0% (0/20) | 0% (0/20) | 0% (0/20) | 0% (0/20) | 0% (0/20) | 0% (0/20) |
| P-value | *nc* | *ref* | *nc* | *nc* | *ref* | *nc* |

## DISCUSSION

To our knowledge, this is the first clinical trial designed to test the gametocytocidal and transmission-blocking properties of the triple artemisinin-based combination therapy artemether-lumefantrine-amodiaquine with and without primaquine and of artesunate-amodiaquine with and without primaquine. Within 48 hours of treatment, transmission was greatly reduced in the artemether-lumefantrine and artemether-lumefantrine-amodiaquine groups, and completely annulled in both treatment groups with primaquine. In contrast, transmission to mosquitoes continued in a minority of individuals until day 28 after treatment with artesunate-amodiaquine alone.

Calls for malaria eradication and the emergence and spread of drug resistance have reinforced the need to assess the effects of antimalarial drugs on gametocytes and their infectiousness (22,23). The addition of a second partner drug to ACTs could significantly delay the emergence and spread of artemisinin resistance and treatment failure. Lumefantrine and amodiaquine provide mutual protection against resistance development, and deployment of the TACT artemether-lumefantrine-amodiaquine is expected to extend the useful lives of artemisinin derivatives and both partner drugs (6). This study was not designed to investigate the clinical efficacy of TACT, nor had the study site recorded any partial artemisinin resistance at the time of the study. We found that both treatments with artemether-lumefantrine; alone and with amodiaquine, greatly reduced transmission by day 2 after treatment. The addition of a single low-dose primaquine only marginally enhanced this transmission-blocking effect. Gametocyte densities minimally differed between artemether-lumefantrine and artemether-lumefantrine-amodiaquine, but we observed a near complete clearance of gametocytes by day 7 in the group with an added single low-dose of primaquine. These observations align with recent data indicating that artemether-lumefantrine has potent transmission-blocking effects (14).

Artesunate-amodiaquine is the first-line treatment for uncomplicated *P. falciparum* malaria in many countries, yet its effect on mature gametocytes and transmission were unclear. In line with previous studies (10,11), we found that gametocyte carriage persisted in all individuals treated with artesunate– amodiaquine until the end of follow-up (day 28) and three (16%) of 19 individuals (three (18%) of 17 individuals infectious at baseline) were still infectious to mosquitoes 14 days after initiation of treatment, with one individual remaining infectious until the end of follow-up (day 28). Moreover, one individual was infectious on day 2 but not at baseline, and one individual was infectious on day 7, but not at baseline or day 2. Whilst there is an inherent stochastic element in transmission to mosquitoes and observing no infected mosquitoes at one timepoint therefore does not rule out (low levels of) infectivity, this pattern suggests a possible role for differences in the drug susceptibility or exposure at different gametocyte developmental stages i.e. immature gametocytes may be released from sequestration in the bone marrow or spleen after treatment. The addition of a single low-dose of primaquine resulted in an enhanced clearance of gametocytes and achieved in a near-total reduction of transmission potential within 48 hours.

The exploratory assay of magnetic gametocyte enrichment showed that the percentage of infectious individuals increased at day 2 after treatment start in all groups except for the artemether-lumefantrine-amodiaquine plus primaquine group. This suggests that in all ACT-only groups, the initial lack of infectivity is due to low gametocyte densities or sex ratio distortion rather than the sterilization of either gametocyte sex. In the primaquine groups, the enrichment results were contradictory: the addition of primaquine to artemether-lumefantrine-amodiaquine blocked all transmission at day 2 even after gametocyte enrichment, suggestive of primaquine sterilizing gametocytes before reducing their numbers significantly (24). Conversely, in the artesunate-amodiaquine plus primaquine group, two individuals who were not infectious in standard feeding assays became infectious after gametocyte concentration. Although these are very small sample sizes and other factors such as variations in primaquine concentration have not been measured, this may indicate varying primaquine efficacy with different artemisinin therapies. Of relevance is that in the process of gametocyte enrichment, human plasma is replaced by malaria-naïve serum, thereby removing drugs that may affect parasite development upon mosquito ingestion.

The emergence of transmission after treatment that was observed in the artesunate-amodiaquine plus primaquine group treatment group in standard feeding assays, has been seen previously after artemisinin (14) and non-artemisinin treatments (14,25). We hypothesise that artesunate-amodiaquine may have lower efficacy on or exposure to immature, developing gametocytes than artemether-lumefantrine, and that those gametocytes released from sequestration after treatment would be unaffected by primaquine’s active metabolites which only circulate for a matter of hours. In addition, it is important to consider that, although artemether-lumefantrine-amodiaquine without primaquine prevents nearly all mosquito infections within 48 hours, gametocytes in *PfKelch13* mutant infections may preferentially survive artemisinin exposure and infect mosquitoes (26). Our data therefore provides strong evidence that the addition of a single low-dose primaquine to artemether-lumefantrine-amodiaquine may be valuable to limit the spread of artemisinin partial resistance in Africa, and supports the suggestion from the WHO malaria policy and advisory group to expand the focus on reducing parasite transmission with a single low-dose of primaquine in areas where partial resistance has been detected (27).

Previous studies reported a higher frequency of side effects with the combination of partner drugs lumefantrine and amodiaquine, compared to lumefantrine alone (7,8), including vomiting, nausea and vertigo, and mild bradycardia. We did not see an increase in vomiting or nausea, and only a slight increase in vertigo related to the drug treatment from one adverse event in the artemether-lumefantrine group, to three and six in the artemether-lumefantrine-amodiaquine groups with and without primaquine, respectively. Overall, all drug regimens were well tolerated, and no instances of cardiac adverse events or severe side effects were reported.

Our study had some limitations. For instance, we assessed a large number of secondary analysis and interpretation therefore requires caution due to issues of multiple testing. In addition, we recruited individuals carrying high densities of gametocytes, consistent with previous studies with similar outcomes and at the same study site (15–18). This allowed us to collect robust data on post-treatment transmissibility but does not represent the average gametocyte-infected individual. Consequently, our estimates of persistence of transmissible gametocytes primarily demonstrate the impact of antimalarial drugs on the transmission potential stemming from a comparatively small subset of highly infectious individuals; though these would be the most important group for the drug regimens to work in. Lastly, it could be argued that the public health significance of these study findings needs to be validated through community trials focused on transmission outcomes. Mass administrations of primaquine or other gametocytocidal compounds (for example, alongside seasonal malaria chemoprophylaxis) may be necessary to achieve reductions in transmission at the community level (28). Conversely, given the negligible cost of primaquine, absence of safety concerns and lack of obvious alternative, there is a compelling argument to add primaquine to slow the transmission of drug resistant parasites.

In conclusion, our findings show that artemether-lumefantrine-amodiaquine can prevent nearly all mosquito infections, but reveal considerable post-treatment transmission after artesunate-amodiaquine. The addition of a single low-dose of primaquine is a safe and effective addition to artemether-lumefantrine-amodiaquine and artesunate-amodiaquine for blocking *P. falciparum* transmission. Enriching the gametocyte content of mosquito blood meals in transmission assays reveals differences in transmission blocking mechanisms for schizonticides and primaquine and strengthens the argument for the addition of a single-low dose of primaquine to block the transmission of artemisinin resistant gametocytes.

### Funding

This work was supported by the Bill and Melinda Gates Foundation (#INV-005735 and #INV-002098). WS is supported by a Wellcome Trust fellowship (218676/Z/19/Z/WT). LNV is supported by a Biotechnology and Biological Sciences Research Council LIDo Ph.D. studentship (BB/T008709/1).

### Contributors

LNV, WS, AM, MJS, CD, and ADi conceived the study and developed the study protocol. AM, WS, LNV, MJS, KS, YSi, SMN, OMD, RSD, MD, SOM, AY, AS, SK, SS, ADe, YD, SFT, CD, and Adi implemented the trial. KL performed molecular analyses. LNV, AM and WS verified the raw data. LNV and WS analysed the data. LNV, AM, WS, MJS, RtH, CD, and ADi wrote the first draft of the manuscript. All authors had full access to all the data in the study and accept responsibility for the decision to submit for publication. All authors read and approved the final manuscript.

### Data sharing

Anonymised data reported in the manuscript will be made available to investigators who provide a methodologically sound proposal to the corresponding author. The study protocol is available in the supplementary information. The data from this trial will be made accessible on the Clinical Epidemiology Database Resources website (https://clinepidb.org).

## Supporting information

Study protocol

## Acknowledgments

Artesunate-amodiaquine, artemether-lumefantrine, and amodiaquine tablets were kindly donated by Guilin Pharmaceutical Co, Shanghai, China. Primaquine tablets were kindly donated by ACE Pharmaceuticals, Zeewolde, The Netherlands. We thank the local safety monitor, members of the data safety and monitoring board for their oversight, and all MRTC study staff for their assistance. Finally, we wish to thank the study participants and the population of Ouélessébougou, Mali, for their cooperation.

