## Supplementary appendix for "Artemether-lumefantrine-amodiaquine or artesunate-amodiaquine combined with single low-dose primaquine to reduce *Plasmodium falciparum* malaria transmission in Ouélessébougou, Mali: a five-arm, phase 2, single-blind, randomised clinical trial"

### Supplementary Information 1. Antimalarial treatment dosing

1. Artemether-Lumefantrine (AL)

Participants in the AL, AL-AQ or AL-AQ+PQ groups were treated with standard doses of AL (Guilin Pharmaceutical, Shanghai, China) from day 0-2. Tablets containing 20 mg artemether and 120 mg lumefantrine will be administered per manufacturer guidelines as shown below:

| **Bodyweight (kg)** | **20/120 mg tablet** | | |
| --- | --- | --- | --- |
|  | **D1** | **D2** | **D3** |
| 5 to < 15 kg | 1 disp tab x 2 | 1 disp tab x 2 | 1 disp tab x 2 |
| 15 to < 25 kg | 2 disp tab x 2 | 2 disp tab x 2 | 2 disp tab x 2 |
| 25 to < 35 kg | 3 tab x 2 | 3 tab x 2 | 3 tab x 2 |
| ≥ 35 kg | 4 tab x 2 | 4 tab x 2 | 4 tab x 2 |

2. Primaquine (PQ)

Participants in the AL-AQ+PQ and AS-AQ+PQ groups received PQ (ACE Pharmaceuticals, Zeewolde, The Netherlands) at a single low dose of 0.25mg/kg as is currently recommended by the World Health Organization. The single dose of PQ was given on day 0 together with the first dose of AL or ASAQ, administered in an aqueous solution, according to a standard operating procedure (SOP) provided Sanofi as previously done at the study site when PQ was combined with DP, PA or AL (1–3).

3. Amodiaquine (AQ)

Participants in the AL-AQ and AL-AQ+PQ groups were given AQ as tablets of 153 mg (Guilin Pharmaceutical, Shanghai, China). The weight-based treatment schedule as shown below aims for a dosage of approximately 10 mg (7.7-15.3mg)/kg/day, given once or twice daily (together with artemether–lumefantrine) for three days:

| **Bodyweight (kg)** | **153 mg tablet** | | | | | |
| --- | --- | --- | --- | --- | --- | --- |
|  | **D1 (0hr)** | **D1 (8hr)** | **D2 (24hr)** | **D2 36hr)** | **D3 (48hr)** | **D3 (60hr)** |
| 10 to 19.9 | 1 tab | 0 tab | 1 tab | 0 tab | 1 tab | 0 tab |
| 20 to 29.9 | 1 tab | 1 tab | 1 tab | 1 tab | 1 tab | 1 tab |
| 30 to 54.9 | 2 tab | 1 tab | 2 tab | 1 tab | 2 tab | 1 tab |
| 55 to 80 | 3 tab | 2 tab | 3 tab | 2 tab | 3 tab | 2 tab |

4. Artesunate-Amodiaquine (AS-AQ)

Participants in the AS-AQ and AS-AQ+PQ groups received fixed-dose combination tablets containing 50mg/135 mg or 100mg/270 mg of artesunate/amodiaquine (Guilin Pharmaceutical, Shanghai, China). Tablets were administered according to manufacturer guidelines, as shown below:

| **Weight** | **Tablets** | **D1** | **D2** | **D3** |
| --- | --- | --- | --- | --- |
| 9 to < 18 kg | 50 mg AS/135 mg AQ base | 1 tab | 1 tab | 1 tab |
| 18 to < 36 kg | 100 mg AS/270 mg AQ base | 1 tab | 1 tab | 1 tab |
|  | blister pack of 3 tab |  |  |  |
| ≥ 36 kg | 100 mg AS/270 mg AQ base | 2 tab | 2 tab | 2 tab |
|  | blister pack of 6 tab |  |  |  |

### Supplementary Figure 1. Schematic representation of sample collection and analysis pipeline

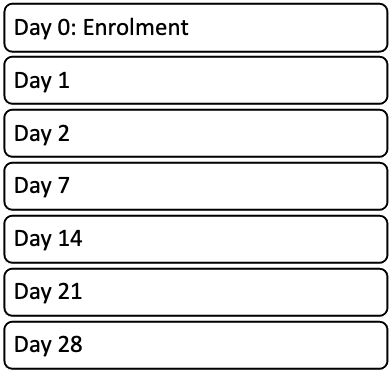

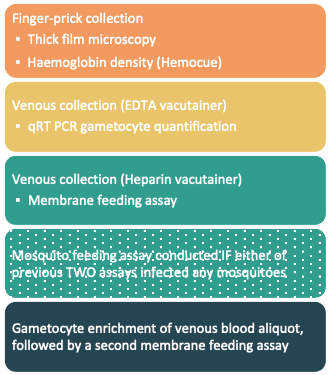

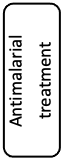

Day 0: Enrolment

Day 1

Day 2

Day 7

Day 14

Day 21

Day 28

Finger-prick collection ▪ Thick film microscopy ▪ Haemoglobin density (Hemocue)

Venous collection (EDTA vacutainer) ▪ qRT PCR gametocyte quantification

Venous collection (Heparin vacutainer) ▪ Membrane feeding assay

Mosquito feeding assay conducted IF either of previous TWO assays infected any mosquitoes

Gametocyte enrichment of venous blood aliquot, followed by a second membrane feeding assay

Antimalarial treatment

### Supplementary Table 1. Primer sequences and qPCR conditions for PfMGET CCp4 assay

**PfMGET Primer/Probe Sequences**

| **Primers** | **Sequence** |
| --- | --- |
| Primer-FW (5’-3’) | CGGTCCAAATATAAAAATCCTG |
| Primer-RV (5’-3’) | TGTG TAACG TATG ATTCATTTTC |
| Probe (5’-3’) | FAM-CAGCTCCAG CATTAAAACAC-BHQ1 |

**CCp4** **Primer/Probe Sequences**

| **Primers** | **Sequence** |
| --- | --- |
| Primer-FW (5’-3’) | CACATGAATATGAGAATAAAATTG |
| Primer-RV (5’-3’) | TAGGCGAACATGTGGAAAG |
| Probe (5’-3’) | TexasRed-AGCAACAACGGTATGTGCCTTAAAACG-BHQ2 |

Male and female gametocyte quantification was performed as described previously, using a multiplex RT-qPCR assay (4). Assays were run using commercial RT-qPCR mixes (Luna® Universal Probe One-Step RT-qPCR Kit, New England Biolabs, Ipswich, MA, USA). FW = Forward primer. RV = Reverse primer.

### Supplementary Table 2. Infectivity to mosquitoes for individuals infectious at baseline

| Day of follow-up | Treatment arm | Infectious individuals  % (n/N) * | p-value^¥^ | p-value^†^ | Mosquito infection rate Median % (IQR) ** | p-value^¥^ | p-value^†^ | Oocyst density Median (IQR) *** | p-value^¥^ | p-value^†^ |
| --- | --- | --- | --- | --- | --- | --- | --- | --- | --- | --- |
| **Day 0** | **Overall** | 61% (61/100) | · | · | 7·3% (3·2-23.5) | · | · | 1·3 (1-3·0) | · | · |
|  | **AL** | 55% (11/20) | *ref* | *ref* | 4·5% (3·3-44·1) | *ref* | *ref* | 1·7 (1-4·8) | *ref* | *ref* |
|  | **AL-AQ** | 55% (11/20) | *ref* | 1·000 | 10·9% (3·3-32·3) | *ref* | 0·646 | 1·3 (1-3·5) | *ref* | 0·756 |
|  | **AL-AQ+PQ** | 55% (11/20) | *ref* | 1·000 | 4·1% (2·1-8·8) | *ref* | 0·045 | 1·2 (1-2·5) | *ref* | 0·535 |
|  | **AS-AQ** | 85% (17/20) | *ref* | *ref* | 7·3% (1·9-23·5) | *ref* | *ref* | 1·3 (1-2·3) | *ref* | *ref* |
|  | **AS-AQ+PQ** | 55% (11/20) | *ref* | 0·082 | 9·3% (1·8-36·2) | *ref* | 0·370 | 1·7 (1-3·9) | *ref* | 0·716 |
| **Day 2** | **AL** | 10% (2/20) | 0·003 | *ref* | 0% (0-0) | 0·0033 | *ref* | 1·0 (1-1) | 0·1797 | *ref* |
|  | **AL-AQ** | 10·5% (2/19) | 0·004 | 0·678 | 0% (0-0) | 0·0033 | 0·523 | 1·0 (1-1) | 0·1797 | *nc* |
|  | **AL-AQ+PQ** | 0% (0/19) | 0·0001 | 0·256 | 0% (0-0) | 0·0033 | 0·788 | *nc* | *nc* | *nc* |
|  | **AS-AQ** | 75% (15/20) | 0·347 | *ref* | 5% (1·5-9·7) | 0·6192 | *ref* | 1·2 (1-1·8) | 0·8744 | *ref* |
|  | **AS-AQ+PQ** | 0% (0/20) | <0·0001 | <0·0001 | 0% (0-0) | 0·0033 | 0·006 | *nc* | *nc* | *nc* |
| **Day 7** | **AL** | 0% (0/20) | <0·0001 | *ref* | 0% (0-0) | 0·0033 | *ref* | *nc* | *nc* | *ref* |
|  | **AL-AQ** | 0% (0/18) | <0·0001 | *nc* | 0% (0-0) | 0·0033 | *nc* | *nc* | *nc* | *nc* |
|  | **AL-AQ+PQ** | 0% (0/19) | <0·0001 | *nc* | 0% (0-0) | 0·0033 | *nc* | *nc* | *nc* | *nc* |
|  | **AS-AQ** | 35% (7/20) | 0·002 | *ref* | 0% (0-6·2) | 0·001 | *ref* | 2·0 (1·6-2·2) | 0·3991 | *ref* |
|  | **AS-AQ+PQ** | 0% (0/20) | <0·0001 | 0·004 | 0% (0-0) | 0·0033 | 0·045 | *nc* | *nc* | *nc* |
| **Day 14** | **AL** | 0% (0/20) | <0·0001 | *ref* | 0% (0-0) | 0·0033 | *ref* | *nc* | *nc* | *ref* |
|  | **AL-AQ** | 0% (0/18) | <0·0001 | *nc* | 0% (0-0) | 0·0033 | *nc* | *nc* | *nc* | *nc* |
|  | **AL-AQ+PQ** | 0% (0/19) | <0·0001 | *nc* | 0% (0-0) | 0·0033 | *nc* | *nc* | *nc* | *nc* |
|  | **AS-AQ** | 15·8% (3/19) | <0·0001 | *ref* | 0% (0-0) | 0·0004 | *ref* | 1 (1-11·9) | 0·1088 | *ref* |
|  | **AS-AQ+PQ** | 0% (0/20) | <0·0001 | 0·106 | 0% (0-0) | 0·0033 | 0·146 | *nc* | *nc* | *nc* |
| **Day 21** | **AL** | 0% (0/20) | <0·0001 | *ref* | 0% (0-0) | *nc* | *ref* | *nc* | *nc* | *ref* |
|  | **AL-AQ** | 0% (0/18) | <0·0001 | *nc* | 0% (0-0) | *nc* | *nc* | *nc* | *nc* | *nc* |
|  | **AL-AQ+PQ** | 0% (0/18) | <0·0001 | *nc* | 0% (0-0) | *nc* | *nc* | *nc* | *nc* | *nc* |
|  | **AS-AQ** | 5·3% (1/19) | <0·0001 | *ref* | 0% (0-0) | 0·0277 | *ref* | 5 (5-5) | *nc* | *ref* |
|  | **AS-AQ+PQ** | 0% (0/20) | <0·0001 | *nc* | 0% (0-0) | *nc* | *nc* | *nc* | *nc* | *nc* |
| **Day 28** | **AL** | 0% (0/20) | <0·0001 | *ref* | 0% (0-0) | *nc* | *ref* | *nc* | *nc* | *ref* |
|  | **AL-AQ** | 0% (0/18) | <0·0001 | *nc* | 0% (0-0) | *nc* | *nc* | *nc* | *nc* | *nc* |
|  | **AL-AQ+PQ** | 0% (0/19) | <0·0001 | *nc* | 0% (0-0) | *nc* | *nc* | *nc* | *nc* | *nc* |
|  | **AS-AQ** | 5·3% (1/19) | <0·0001 | *ref* | 0% (0-10·3) | 0·1088 | *ref* | 1 (1-1) | *nc* | *ref* |
|  | **AS-AQ+PQ** | 0% (0/20) | <0·0001 | *nc* | 0% (0-0) | *nc* | *nc* | *nc* | *nc* | *nc* |

*Percentage of infectious individuals. Individuals were classed as infectious if direct membrane feeding assays (DMFA) resulted in at least one mosquito with any number of oocysts. Mosquito infection measures (mosquito infection rate and oocyst density) are presented for all participants who were infectious at baseline, and oocyst densities are from all infected mosquitoes **Mosquito infection rate is the median percentage of mosquitoes infected by each participant, where for each participant mosquito infection rate the number of mosquitoes infected as a percentage of all mosquitoes surviving to dissection. ***The average oocyst density for each participant was calculated as the mean number of oocysts in infected mosquitoes (i.e., with at least one oocyst). The value presented in the table is the median of all individuals’ average oocyst intensities (a composite figure of all oocysts/all infected mosquitoes is not statistically valid). P-value¥ = Within group comparison. P-value† = Between group comparison (artemether–lumefantrine vs artemether–lumefantrine-amodiaquine and artemether–lumefantrine-amodiaquine plus primaquine, artesunate-amodiaquine vs artesunate-amodiaquine plus primaquine). nc = not calculable, no positive observations. · = not tested, ref = reference group. AL = artemether-lumefantrine; AL-AQ = artemether-lumefantrine-amodiaquine; AL-AQ+PQ = artemether-lumefantrine-amodiaquine plus primaquine; AS-AQ = artesunate-amodiaquine; AS-AQ+PQ = artesunate-amodiaquine plus primaquine

**Supplementary Table 3.** **Infectivity to mosquitoes for all individuals**

| Day of follow-up | Treatment arm | Mosquito infection rate  Median % (IQR) * | p-value^¥^ | p-value^†^ | Oocyst density Median (IQR) ** | p-value^¥^ | p-value^†^ | Median % reduction in mosquito infection rate (IQR) ******* | p-value^¥^ | p-value^†^ |
| --- | --- | --- | --- | --- | --- | --- | --- | --- | --- | --- |
| **Day 0** | **Overall** | 2·0% (0-9·4) | · | · | 1·3 (1-3·0) | · | · | · | · | · |
|  | **AL** | 3·0% (0-13·6) | *ref* | *ref* | 1·7 (1-4·8) | *ref* | *ref* | · | *ref* | · |
|  | **AL-AQ** | 2·5% (0-11·2) | *ref* | 0·686 | 1·3 (1-3·5) | *ref* | 0·7563 | · | *ref* | · |
|  | **AL-AQ+PQ** | 1·6% (0-4·8) | *ref* | 0·071 | 1·2 (1-2·5) | *ref* | 0·5349 | · | *ref* | · |
|  | **AS-AQ** | 3·9% (1·6-19·8) | *ref* | *ref* | 1·3 (1-2·3) | *ref* | *ref* | · | *ref* | · |
|  | **AS-AQ+PQ** | 1·5% (0-9·4) | *ref* | 0·955 | 1·7 (1-3·9) | *ref* | 0·7158 | · | *ref* | · |
| **Day 2** | **AL** | 0% (0-0) | 0·0012 | *ref* | 1·0 (1-1) | 0·1797 | *ref* | 88·19 (0-100) | 0·001 | *ref* |
|  | **AL-AQ** | 0% (0-0) | 0·0012 | 0·513 | 1·0 (1-1) | 0·1797 | *nc* | 95·30 (0-100) | 0·001 | 0·8644 |
|  | **AL-AQ+PQ** | 0% (0-0) | 0·0012 | 0·750 | *nc* | *nc* | *nc* | 100 (0-100) | 0·0009 | 0·6003 |
|  | **AS-AQ** | 2·3% (0·7-8·2) | 0·614 | *ref* | 1·2 (1-1·8) | 0·8744 | *ref* | 7·40 (-9·40-72·91) | 0·1901 | *ref* |
|  | **AS-AQ+PQ** | 0% (0-0) | 0·0012 | 0·006 | *nc* | *nc* | *nc* | 100 (0-100) | 0·0009 | 0·0465 |
| **Day 7** | **AL** | 0% (0-0) | 0·0012 | *ref* | *nc* | *nc* | *ref* | 100 (0-100) | 0·0009 | *ref* |
|  | **AL-AQ** | 0% (0-0) | 0·0013 | *nc* | *nc* | *nc* | *nc* | 100 (0-100) | 0·0009 | 0·707 |
|  | **AL-AQ+PQ** | 0% (0-0) | 0·0012 | *nc* | *nc* | *nc* | *nc* | 100 (0-100) | 0·0009 | 0·8573 |
|  | **AS-AQ** | 0% (0-4) | 0·0011 | *ref* | 2·0 (1·0-2·2) | 0·3991 | *ref* | 100 (32·38-100) | 0·0018 | *ref* |
|  | **AS-AQ+PQ** | 0% (0-0) | 0·0012 | 0·070 | *nc* | *nc* | *nc* | 100 (0-100) | 0·0009 | 0·5878 |
| **Day 14** | **AL** | 0% (0-0) | 0·0012 | *ref* | *nc* | *nc* | *ref* | 100 (0-100) | 0·0009 | *ref* |
|  | **AL-AQ** | 0% (0-0) | 0·0013 | *nc* | *nc* | *nc* | *nc* | 100 (0-100) | 0·0009 | 0·707 |
|  | **AL-AQ+PQ** | 0% (0-0) | 0·0012 | *nc* | *nc* | *nc* | *nc* | 100 (0-100) | 0·0009 | 0·8573 |
|  | **AS-AQ** | 0% (0-0) | 0·0002 | *ref* | 1 (1-11·9) | 0·1088 | *ref* | 100 (90·23-100) | 0·0001 | *ref* |
|  | **AS-AQ+PQ** | 0% (0-0) | 0·0012 | 0·236 | *nc* | *nc* | *nc* | 100 (0-100) | 0·0009 | 0·2022 |
| **Day 21** | **AL** | 0% (0-0) | *nc* | *ref* | *nc* | *nc* | *ref* | *nc** | *nc** | *ref* |
|  | **AL-AQ** | 0% (0-0) | *nc* | *nc* | *nc* | *nc* | *nc* | *nc** | *nc** | *nc** |
|  | **AL-AQ+PQ** | 0% (0-0) | *nc* | *nc* | *nc* | *nc* | *nc* | *nc** | *nc** | *nc** |
|  | **AS-AQ** | 0% (0-0) | 0·022 | *ref* | 5 (5-5) | *nc* | *ref* | 100 (77·24-100) | 0·0174 | *ref* |
|  | **AS-AQ+PQ** | 0% (0-0) | *nc* | *nc* | *nc* | *nc* | *nc* | *nc** | *nc** | *nc** |
| **Day 28** | **AL** | 0% (0-0) | *nc* | *ref* | *nc* | *nc* | *ref* | *nc** | *nc** | *ref* |
|  | **AL-AQ** | 0% (0-0) | *nc* | *nc* | *nc* | *nc* | *nc* | *nc** | *nc** | *nc** |
|  | **AL-AQ+PQ** | 0% (0-0) | *nc* | *nc* | *nc* | *nc* | *nc* | *nc** | *nc** | *nc** |
|  | **AS-AQ** | 0% (0-10·3) | 0·1088 | *ref* | 1 (1-1) | *nc* | *ref* | 100 (83·85-100) | 0·1025 | *ref* |
|  | **AS-AQ+PQ** | 0% (0-0) | *nc* | *nc* | *nc* | *nc* | *nc* | *nc** | *nc** | *nc** |

Mosquito infection measure (mosquito infection rate, oocyst density and reduction in mosquito infection rate) are presented for all individuals regardless of baseline infectivity. *Mosquito infection rate is the median percentage of mosquitoes infected by each participant, where for each participant mosquito infection rate the number of mosquitoes infected as a percentage of all mosquitoes surviving to dissection. **The average oocyst density for each participant was calculated as the mean number of oocysts in infected mosquitoes (i.e., with at least one oocyst). The value presented in the table is the median of all individuals’ average oocyst intensities (a composite figure of all oocysts/all infected mosquitoes is not statistically valid). *** Median within-person (relative to baseline) reduction in mosquito infection, including individuals not infectious at baseline. P-value¥ = Within group comparison. P-value† = Between group comparison (artemether–lumefantrine vs artemether–lumefantrine-amodiaquine and artemether–lumefantrine-amodiaquine plus primaquine, artesunate-amodiaquine vs artesunate-amodiaquine plus primaquine). nc = not calculable, no positive observations. nc* = not calculable, one or zero individuals remained infectious and participated in mosquito feeding. · = not tested, ref = reference group. AL = artemether-lumefantrine; AL-AQ = artemether-lumefantrine-amodiaquine; AL-AQ+PQ = artemether-lumefantrine-amodiaquine plus primaquine; AS-AQ = artesunate-amodiaquine; AS-AQ+PQ = artesunate-amodiaquine plus primaquine

**Supplementary Figure 2. Difference in mosquito infection rate and person infectivity before and after gametocyte enrichment**

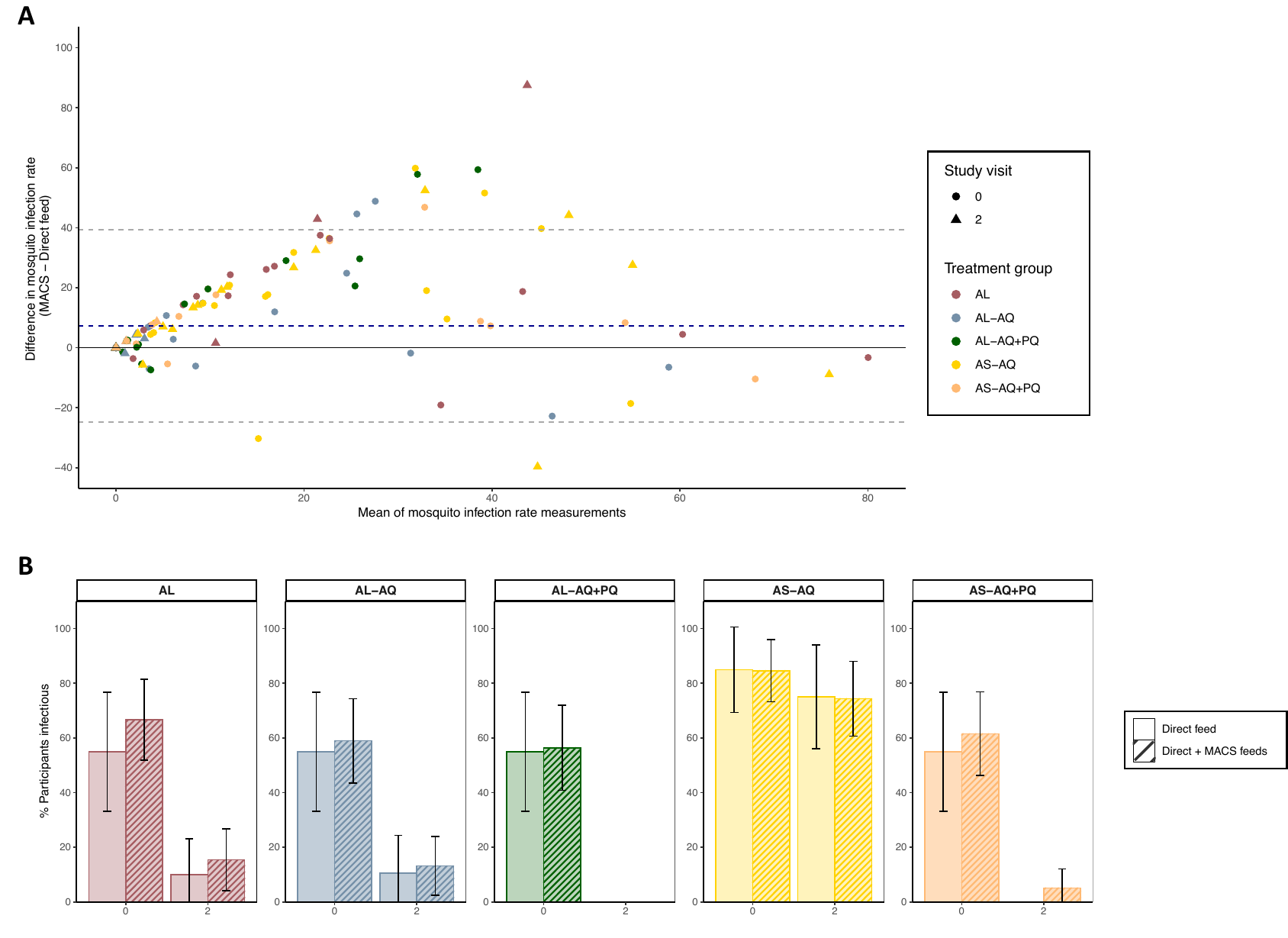

**A.** Difference in mosquito infection rate comparing direct feeds to gametocyte enriched feeds (after MACS). Colour indicates treatment group and shape indicates the study visit. Blue dashed line indicates the mean of the difference between measurements across all samples. Grey dashed lines represent ± 1·96 standard deviations (SD) of the mean. **B.** Person infectivity by direct membrane feeding assay compared to direct membrane feeding assay boosted by gametocyte enrichment at baseline and day 2 post-treatment. AL = artemether-lumefantrine; AL-AQ = artemether-lumefantrine-amodiaquine; AL-AQ+PQ = artemether-lumefantrine-amodiaquine plus primaquine; AS-AQ = artesunate-amodiaquine; AS-AQ+PQ = artesunate-amodiaquine plus primaquine

### Supplementary Table 4. Participant infectivity after gametocyte enrichment.

|  | **Direct membrane feed** | **Direct and gametocyte enriched membrane feeds** | | |
| --- | --- | --- | --- | --- |
|  | **Infectious individuals % (n/N)** | **Infectious individuals % (n/N)** | **p-value*^¥^*** | **p-value†** |
| **Day 0** | | | | |
| **AL** | 55% (11/20) | 85% (17/20) | 0·041 | *ref* |
| **AL-AQ** | 55% (11/20) | 65% (13/20) | 0·374 | 0·273 |
| **AL-AQ+PQ** | 55% (11/20) | 75% (15/20) | 0·160 | 0·695 |
| **AS-AQ** | 85% (17/20) | 90% (18/20) | 0·500 | *ref* |
| **AS-AQ+PQ** | 55% (11/20) | 70% (14/20) | 0·257 | 0·235 |
| **Day 2** | | | | |
| **AL** | 10% (2/20) | 20% (5/20) | 0·204 | *ref* |
| **AL-AQ** | 10·5% (2/19) | 21% (4/19) | 0·330 | 0·535 |
| **AL-AQ+PQ** | 0% (0/19) | 0% (0/19) | nc | 0·027 |
| **AS-AQ** | 75% (15/20) | 80% (16/20) | 0·500 | *ref* |
| **AS-AQ+PQ** | 0% (0/20) | 10% (2/20) | 0·244 | <0·0001 |

Participant infectivity after direct membrane feeding assay and gametocyte enrichment-boosted direct membrane feeding assays at baseline and day 2 after treatment initiation. ^¥^Within-group comparison (direct feed as reference). †Between artemisinin-based combination therapy matched group comparison (i.e., artemether–lumefantrine vs artemether–lumefantrine-amodiaquine and artemether–lumefantrine-amodiaquine plus primaquine, artesunate-amodiaquine vs artesunate-amodiaquine plus primaquine) by Fishers exact test. Nc = not calculable, ref = reference group. AL = artemether-lumefantrine; AL-AQ = artemether-lumefantrine-amodiaquine; AL-AQ+PQ = artemether-lumefantrine-amodiaquine plus primaquine; AS-AQ = artesunate-amodiaquine; AS-AQ+PQ = artesunate-amodiaquine plus primaquine

### Supplementary Table 5. Asexual parasite density by microscopy

| **Day of follow-up** | **Treatment arm** | **Median asexual parasites/µL (IQR)** | **p-value*^¥^*** | **p-value†** | **Prevalence**  **n/N (%)** | **p-value*^¥^*** | **p-value†** |
| --- | --- | --- | --- | --- | --- | --- | --- |
| **Day 0** | **Overall** | 0·00 (0·00-241·99) | · | · | 37% (37/96) | · | · |
|  | **AL** | 37·9 (0·0-300·0) | *ref* | *ref* | 50% (10/20) | *ref* | *ref* |
|  | **AL-AQ** | 0·0 (0·0-79·8) | *ref* | 0·1193 | 30% (6/20) | *ref* | 0·167 |
|  | **AL-AQ+PQ** | 0·0 (0·0-37·6) | *ref* | 0·0871 | 25% (5/20) | *ref* | 0·095 |
|  | **AS-AQ** | 0·0 (0·0-1654·9) | *ref* | *ref* | 40% (8/20) | *ref* | *ref* |
|  | **AS-AQ+PQ** | 0·0 (0·0-720·0) | *ref* | 0·5824 | 40% (8/20) | *ref* | 0·626 |
| **Day 2** | **AL** | 0·0 (0·0-0·0) | 0·0031 | *ref* | 5% (1/20) | 0·002 | *ref* |
|  | **AL-AQ** | 0·0 (0·0-0·0) | 0·0148 | 0·3297 | 0% (0/19) | 0·012 | 0·513 |
|  | **AL-AQ+PQ** | 0·0 (0·0-0·0) | 0·0459 | 0·3297 | 0% (0/19) | 0·027 | 0·513 |
|  | **AS-AQ** | 0·0 (0·0-0·0) | 0·0051 | *ref* | 5% (1/20) | 0·010 | *ref* |
|  | **AS-AQ+PQ** | 0·0 (0·0-0·0) | 0·0051 | 0·3173 | 0% (0/20) | 0·002 | 0·500 |
| **Day 7** | **AL** | 0·0 (0·0-0·0) | 0·0019 | *ref* | 0% (0/20) | 0·0002 | *ref* |
|  | **AL-AQ** | 0·0 (0·0-0·0) | 0·0149 | nc | 0% (0/18) | 0·014 | nc |
|  | **AL-AQ+PQ** | 0·0 (0·0-0·0) | 0·0459 | nc | 0% (0/19) | 0·027 | nc |
|  | **AS-AQ** | 0·0 (0·0-0·0) | 0·0051 | *ref* | 0% (0/20) | 0·002 | *ref* |
|  | **AS-AQ+PQ** | 0·0 (0·0-0·0) | 0·0051 | nc | 0% (0/20) | 0·002 | nc |
| **Day 14** | **AL** | 0·0 (0·0-0·0) | 0·0019 | *ref* | 0% (0/20) | 0·0002 | *ref* |
|  | **AL-AQ** | 0·0 (0·0-0·0) | 0·0149 | nc | 0% (0/18) | 0·014 | nc |
|  | **AL-AQ+PQ** | 0·0 (0·0-0·0) | 0·0459 | nc | 0% (0/19) | 0·027 | nc |
|  | **AS-AQ** | 0·0 (0·0-0·0) | 0·0087 | *ref* | 0% (0/19) | 0·002 | *ref* |
|  | **AS-AQ+PQ** | 0·0 (0·0-0·0) | 0·0051 | nc | 0% (0/20) | 0·002 | nc |
| **Day 21** | **AL** | 0·0 (0·0-0·0) | 0·0019 | *ref* | 0% (0/20) | 0·0002 | *ref* |
|  | **AL-AQ** | 0·0 (0·0-0·0) | 0·0149 | nc | 0% (0/18) | 0·014 | nc |
|  | **AL-AQ+PQ** | 0·0 (0·0-0·0) | 0·0836 | nc | 0% (0/18) | 0·031 | nc |
|  | **AS-AQ** | 0·0 (0·0-0·0) | 0·0087 | *ref* | 0% (0/19) | 0·002 | *ref* |
|  | **AS-AQ+PQ** | 0·0 (0·0-0·0) | 0·0051 | nc | 0% (0/20) | 0·002 | nc |
| **Day 28** | **AL** | 0·0 (0·0-0·0) | 0·0139 | *ref* | 5% (1/20) | 0·002 | *ref* |
|  | **AL-AQ** | 0·0 (0·0-0·0) | 0·0149 | 0·3428 | 0% (0/18) | 0·014 | 0·526 |
|  | **AL-AQ+PQ** | 0·0 (0·0-0·0) | 0·0459 | 0·3297 | 0% (0/19) | 0·027 | 0·513 |
|  | **AS-AQ** | 0·0 (0·0-0·0) | 0·0087 | *ref* | 0% (0/19) | 0·002 | *ref* |
|  | **AS-AQ+PQ** | 0·0 (0·0-0·0) | 0·0051 | nc | 0% (0/20) | 0·002 | nc |

Asexual parasite density (asexual parasites / µL) and prevalence of asexual parasites at all time points, measured by thick film microscopy (counted against 200 WBC). ^¥^Within-group comparisons †Between artemisinin-based combination therapy matched group comparison (i.e., artemether–lumefantrine vs artemether–lumefantrine-amodiaquine and artemether–lumefantrine-amodiaquine plus primaquine, artesunate-amodiaquine vs artesunate-amodiaquine plus primaquine). Nc = not calculable, ref = reference group. AL = artemether-lumefantrine; AL-AQ = artemether-lumefantrine-amodiaquine; AL-AQ+PQ = artemether-lumefantrine-amodiaquine plus primaquine; AS-AQ = artesunate-amodiaquine; AS-AQ+PQ = artesunate-amodiaquine plus primaquine

### Supplementary Figure 3. Gametocyte density and prevalence by gametocyte sex

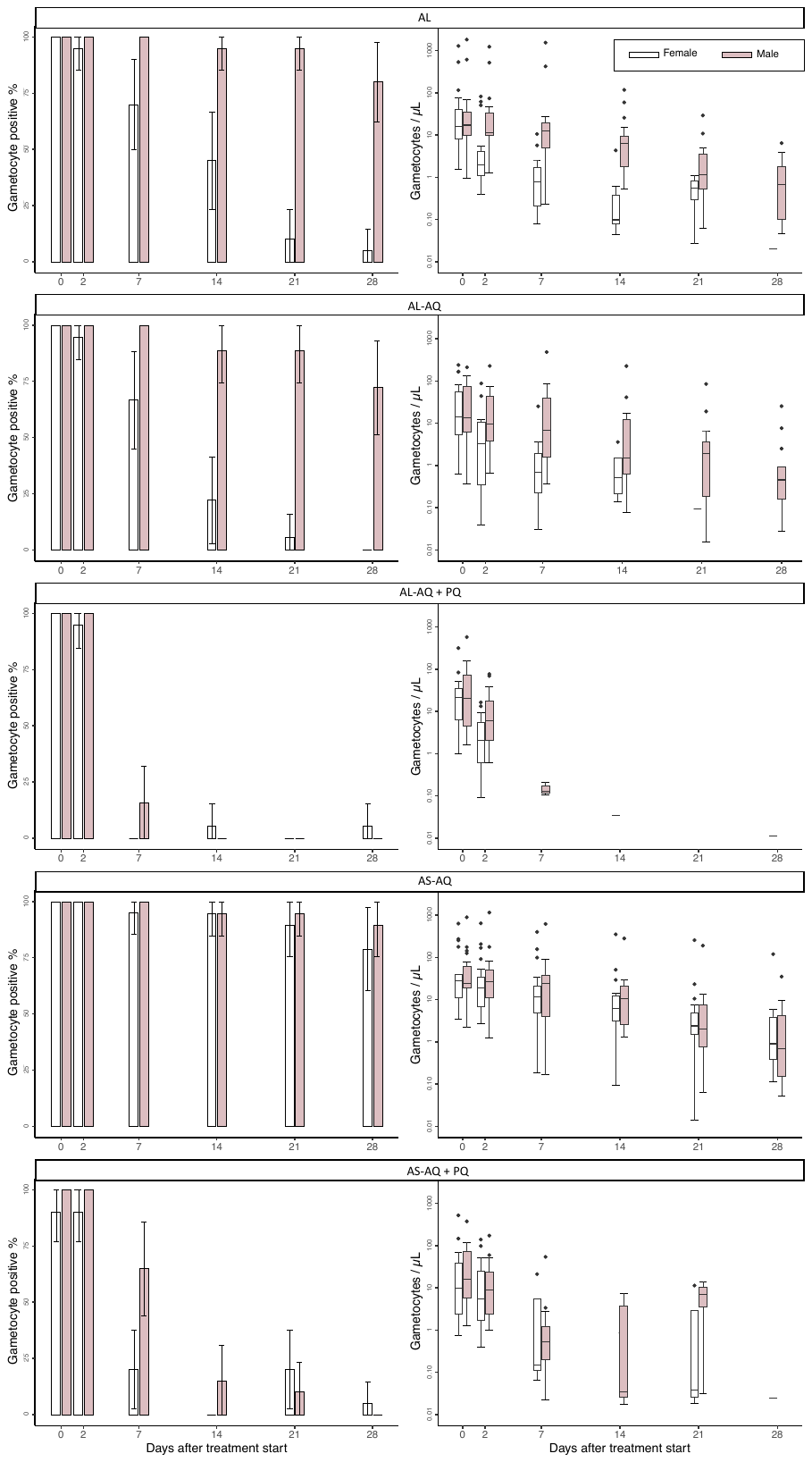

AL = artemether-lumefantrine; AL-AQ = artemether-lumefantrine-amodiaquine; AL-AQ+PQ = artemether-lumefantrine-amodiaquine plus primaquine; AS-AQ = artesunate-amodiaquine; AS-AQ+PQ = artesunate-amodiaquine plus primaquine

### Supplementary Table 6. Total gametocyte density, prevalence and sex ratio

| **Day of follow-up** | **Total gametocytes (CCP4 & PfMGET)** | | | | | | |
| --- | --- | --- | --- | --- | --- | --- | --- |
|  | **Treatment arm** | **Median gametocytes/µL**  **(IQR)** | **p-value** | **Prevalence**  **n/N (%)** | **p-value** | **Proportion male**  **Median (IQR)** | **p-value** |
| **Day 0** | **Overall** | 38·01 (13·55-113·01) | · | 100% (100/100) | · | 0·54 (0·42-0·65) | · |
|  | **AL** | 30·96 (19·90-92·72) | *ref* | 100% (20/20) | *ref* | 0·50 (0·42-0·65) | *ref* |
|  | **AL-AQ** | 28·58 (11·49-130·45) | 0·908 | 100% (20/20) | *nc* | 0·53 (0·42-0·62) | 0·9784 |
|  | **AL-AQ+PQ** | 42·30 (11·79-97·04) | 0·838 | 100% (20/20) | *nc* | 0·52 (0·43-0·70) | 0·3302 |
|  | **AS-AQ** | 52·52 (33·59-128·97) | *ref* | 100% (20/20) | *ref* | 0·52 (0·37-0·64) | *ref* |
|  | **AS-AQ+PQ** | 24·82 (10·74-115·22) | 0·105 | 100% (20/20) | *nc* | 0·63 (0·49-0·68) | 0·1231 |
| **Day 2** | **AL** | 15·43 (10·30-43·69) | *ref* | 100% (20/20) | *ref* | 0·88 (0·77-0·94) | *ref* |
|  | **AL-AQ** | 11·39 (4·12-59·23) | 0·684 | 100% (19/19) | *nc* | 0·82 (0·77-0·92) | 0·4397 |
|  | **AL-AQ+PQ** | 7·77 (3·43-25·28) | 0·066 | 100% (19/19) | *nc* | 0·77 (0·68-0·93) | 0·1559 |
|  | **AS-AQ** | 45·56 (19·18-100·32) | *ref* | 100% (20/20) | *ref* | 0·50 (0·44-0·67) | *ref* |
|  | **AS-AQ+PQ** | 12·64 (5·11-49·64) | 0·011 | 100% (20/20) | *nc* | 0·54 (0·41-0·71) | 0·7251 |
| **Day 7** | **AL** | 13·25 (5·61-21) | *ref* | 100% (20/20) | *ref* | 0·98 (0·91-1) | *ref* |
|  | **AL-AQ** | 6·89 (1·05-49·86) | 0·937 | 100% (18/18) | *nc* | 0·99 (0·93-1) | 0·6775 |
|  | **AL-AQ+PQ** | 0 (0-0) | 0·002 | 15·8% (3/19) | <0·0001 | 1 (1-1) | 0·2389 |
|  | **AS-AQ** | 31·74 (7·27-61·59) | *ref* | 100% (20/20) | *ref* | 0·59 (0·36-0·73) | *ref* |
|  | **AS-AQ+PQ** | 0·17 (0-0·87) | <0·0001 | 70·0% (14/20) | 0·010 | 1 (0·88-1) | 0·0002 |
| **Day 14** | **AL** | 5·61 (1·41-9·30) | *ref* | 95·0% (19/20) | *ref* | 1 (0·98-1) | *ref* |
|  | **AL-AQ** | 1·44 (0·37-11·63) | 0·899 | 88·9% (16/18) | 0·459 | 1 (1-1) | 0·2648 |
|  | **AL-AQ+PQ** | 0 (0-0) | 0·246 | 5·3% (1/19) | <0·0001 | *nc* | *nc* |
|  | **AS-AQ** | 13·65 (4·93-32) | *ref* | 94·7% (18/19) | *ref* | 0·49 (0·38-0·68) | *ref* |
|  | **AS-AQ+PQ** | 0 (0-0) | 0·014 | 15·0% (3/20) | <0·0001 | 1 (1-1) | 0·1003 |
| **Day 21** | **AL** | 1·02 (0·50-3·60) | *ref* | 95·0% (19/20) | *ref* | 1 (1-1) | *ref* |
|  | **AL-AQ** | 1·16 (0·09-3·44) | 0·212 | 88·9% (16/18) | 0·459 | 1 (1-1) | 0·7063 |
|  | **AL-AQ+PQ** | 0 (0-0) | *nc* | 0% (0/18) | <0·0001 | *nc* | *nc* |
|  | **AS-AQ** | 4·25 (0·91-13·39) | *ref* | 94·7% (18/19) | *ref* | 0·47 (0·37-0·72) | *ref* |
|  | **AS-AQ+PQ** | 0 (0-0) | 0·500 | 20·0% (4/20) | <0·0001 | 0·55 (0·55-0·55) | 0·6299 |
| **Day 28** | **AL** | 0·21 (0·06-1·48) | *ref* | 80·0% (16/20) | *ref* | 1 (1-1) | *ref* |
|  | **AL-AQ** | 0·21 (0-0·63) | 0·307 | 72·2% (13/18) | 0·427 | 1 (1-1) | *nc* |
|  | **AL-AQ+PQ** | 0 (0-0) | 0·579 | 5·3% (1/19) | <0·0001 | *nc* | *nc* |
|  | **AS-AQ** | 1·22 (0·22-7·23) | *ref* | 94·7% (18/19) | *ref* | 0·42 (0·27-0·90) | *ref* |
|  | **AS-AQ+PQ** | 0 (0-0) | 0·543 | 5·0% (1/20) | <0·0001 | *nc* | *nc* |

P-values are for differences between artemisinin-based combination therapy matched group comparison (i.e., artemether–lumefantrine vs artemether–lumefantrine-amodiaquine and artemether–lumefantrine-amodiaquine plus primaquine, artesunate-amodiaquine vs artesunate-amodiaquine plus primaquine). Density was compared using regression analyses of log10 transformed density values, with adjustment for baseline densities. Prevalence was compared with one sided Fishers exact tests. For males and females, proportion male is given for participants/time-points with total gametocyte densities of 0·2/µL and over, as described previously (1). For the calculation of gametocyte prevalence, samples were classified as negative for a particular gametocyte sex if the estimated density of in gametocytes of that sex was less than 0·01/μL (i.e. one gametocyte per 100 μL of blood sample). P-value§ = within group comparison. P-value¶ = between group comparison. nc = not calculable, no observations/no observations over the threshold density for analysis, ·= not tested, *ref* = reference group, AL = artemether-lumefantrine; AL-AQ = artemether-lumefantrine-amodiaquine; AL-AQ+PQ = artemether-lumefantrine-amodiaquine plus primaquine; AS-AQ = artesunate-amodiaquine; AS-AQ+PQ = artesunate-amodiaquine plus primaquine.

### Supplementary Table 7. Gametocyte circulation time and area under the curve

|  | **Treatment group** | **Total gametocytes**  **(CCP4 & PfMGET)** | **p-value*** | **Female gametocytes (CCP4)** | **p-value*** | **Male gametocytes (PfMGET)** | **p-value*** | **p-value♂♀** |
| --- | --- | --- | --- | --- | --- | --- | --- | --- |
| **Circulation time**  **Days (95% CI)** | **AL** | 6·13 (5·36-6·90) | *ref* | 3·19 (2·69-3·70) | *ref* | 6·83 (5·95-7·72) | *ref* | <0·0001 |
|  | **AL-AQ** | 6·00 (5·20-6·79) | 0·8071 | 2·75 (2·17-3·32) | 0·2410 | 6·75 (5·82-7·68) | 0·8951 | <0·0001 |
|  | **AL-AQ+PQ** | 2·60 (2·06-3·13) | <0·0001 | 3·27 (2·31-4·24) | 0·8824 | 1·31 (1·00-1·63) | <0·0001 | 0·0006 |
|  | **AS-AQ** | 7·99 (6·70-9·28) | *ref* | 9·07 (7·16-10·98) | *ref* | 7·77 (6·63-8·91) | *ref* | 0·0066 |
|  | **AS-AQ+PQ** | 3·30 (2·79-3·81) | <0·0001 | 4·81 (3·45-6·17) | 0·0005 | 3·43 (2·76-4·11) | <0·0001 | 0·1191 |
| **AUC Median (IQR) gametocytes per uL/day** | **AL** | 9·36 (5·31-21·91) | *ref* | 0·88 (0·61-2·21) | *ref* | 7·79 (3·96-17·55) | *ref* | <0·0001 |
|  | **AL-AQ** | 5·35 (2·21-40·44) | 0·323 | 1·11 (0·33-3·02) | 0·925 | 3·55 (1·54-26·71) | 0·071 | <0·0001 |
|  | **AL-AQ+PQ** | 4·42 (1·22-14·86) | <0·0001 | 1·15 (0·31-2·69) | 0·057 | 2·42 (0·75-10·78) | <0·0001 | <0·0001 |
|  | **AS-AQ** | 26·39 (12·80-51·70) | *ref* | 7·23 (5·03-23·10) | *ref* | 19·64 (4·83-27·47) | *ref* | 0·0337 |
|  | **AS-AQ+PQ** | 6·03 (2·84-18·41) | 0·002 | 1·35 (0·23-5·84) | 0·001 | 3·19 (2·27-11·11) | 0·007 | 0·0003 |

Gametocyte circulation time was calculated using a deterministic compartmental model (5), and is presented as the model estimate (mean days) with 95% CI. Area under the curve (AUC) of gametocyte density per participant over time was calculated using the linear trapezoid method (6), and is presented as the median and IQR of individual AUC values by treatment arm. P-values are for differences in the t-statistic between AL-AQ, AL-AQ+PQ and the AL reference group, and between AS-AQ+PQ and the AS-AQ reference group (*), and for between sexes within treatment groups (♂♀). *Ref* = reference, AL = artemether-lumefantrine; AL-AQ = artemether-lumefantrine-amodiaquine; AL-AQ+PQ = artemether-lumefantrine-amodiaquine plus primaquine; AS-AQ = artesunate-amodiaquine; AS-AQ+PQ = artesunate-amodiaquine plus primaquine.

### Supplementary Table 8. Female (CCP4) and male (PfMGET) gametocyte density and prevalence

| Day of follow-up | Treatment arm | Female gametocytes (CCP4) | | | | Male gametocytes (PfMGET) | | | |
| --- | --- | --- | --- | --- | --- | --- | --- | --- | --- |
|  |  | Median/µL  (IQR) | p-value | Prevalence  n/N (%) | p-value | Median/µL  (IQR) | p-value | Prevalence  n/N (%) | p-value |
| Day 0 | Overall | 15·63 (6·12-39·20) | · | 98% (98/100) | · | 19·72 (8·52-69·39) | · | 100% (100/100) | · |
|  | AL | 16·26 (7·83-47·60) | *ref* | 100% (20/20) | *ref* | 17·19 (9·52-39·56) | *ref* | 100% (20/20) | *ref* |
|  | AL-AQ | 14·07 (4·87-61·79) | 0·890 | 100% (20/20) | *nc* | 13·70 (6·20-77·32) | 0·825 | 100% (20/20) | *nc* |
|  | AL-AQ+PQ | 21·19 (5·73-35·53) | 0·935 | 100% (20/20) | *nc* | 20·54 (4·44-75·63) | 0·637 | 100% (20/20) | *nc* |
|  | AS-AQ | 28·10 (11·26-39·20) | *ref* | 100% (20/20) | *ref* | 24·18 (19·48-66·89) | *ref* | 100% (20/20) | *ref* |
|  | AS-AQ+PQ | 7·47 (1·43-38·49) | 0·090 | 90% (18/20) | 0·244 | 15·98 (5·08-76·78) | 0·234 | 100% (20/20) | *nc* |
| Day 2 | AL | 1·81 (0·98-4·05) | *ref* | 95% (19/20) | *ref* | 11·56 (9·69-35·11) | *ref* | 100% (20/20) | *ref* |
|  | AL-AQ | 2·35 (0·29-10·90) | 0·207 | 94·7% (18/19) | 0·744 | 9·55 (3·60-48·33) | 0·701 | 100% (19/19) | *nc* |
|  | AL-AQ+PQ | 1·95 (0·46-6·23) | 0·880 | 94·7% (18/19) | 0·744 | 6·03 (1·94-18·23) | 0·040 | 100% (19/19) | *nc* |
|  | AS-AQ | 18·61 (6·51-41·24) | *ref* | 100% (20/20) | *ref* | 27·25 (10·53-57·52) | *ref* | 100% (20/20) | *ref* |
|  | AS-AQ+PQ | 4·35 (1·36-22·11) | 0·018 | 90% (18/20) | 0·244 | 8·94 (2·29-28·44) | 0·024 | 100% (20/20) | *nc* |
| Day 7 | AL | 0·26 (0-1·23) | *ref* | 70% (14/20) | *ref* | 12·46 (4·87-20·28) | *ref* | 100% (20/20) | *ref* |
|  | AL-AQ | 0·18 (0-1·17) | 0·566 | 66·7% (12/18) | 0·550 | 6·68 (1·05-46·15) | 1·000 | 100% (18/18) | *nc* |
|  | AL-AQ+PQ | 0 (0-0) | *nc* | 0% (0/19) | <0·0001 | 0 (0-0) | 0·003 | 15·8% (3/19) | <0·0001 |
|  | AS-AQ | 11·07 (4·06-21·03) | *ref* | 95% (19/20) | *ref* | 24·15 (3·78-39·47) | *ref* | 100% (20/20) | *ref* |
|  | AS-AQ+PQ | 0 (0-0) | 0·064 | 20% (4/20) | <0·0001 | 0·17 (0-0·87) | 0·00042 | 65% (13/20) | 0·004 |
| Day 14 | AL | 0 (0-0·10) | *ref* | 45% (9/20) | *ref* | 5·61 (1·41-9·26) | *ref* | 95% (19/20) | *ref* |
|  | AL-AQ | 0 (0-0) | 0·668 | 22·2% (4/18) | 0·128 | 1·44 (0·37-11·38) | 0·908 | 88·9% (16/18) | 0·459 |
|  | AL-AQ+PQ | 0 (0-0) | 0·729 | 5·3% (1/19) | 0·005 | 0 (0-0) | *nc* | 0% (0/19) | <0·0001 |
|  | AS-AQ | 5·61 (2·99-12·68) | *ref* | 94·7% (18/19) | *ref* | 10·23 (1·68-21·99) | *ref* | 94·7% (18/19) | *ref* |
|  | AS-AQ+PQ | 0 (0-0) | *nc* | 0% (0/20) | <0·0001 | 0 (0-0) | 0·115 | 15% (3/20) | <0·0001 |
| Day 21 | AL | 0 (0-0) | *ref* | 10·0% (2/20) | *ref* | 1·02 (0·50-3·6) | *ref* | 95% (19/20) | *ref* |
|  | AL-AQ | 0 (0-0) | *nc* | 5·6% (1/18) | 0·541 | 1·16 (0·09-3·44) | 0·199 | 88·9% (16/18) | 0·459 |
|  | AL-AQ+PQ | 0 (0-0) | *nc* | 0% (0/18) | 0·270 | 0 (0-0) | *nc* | 0% (0/18) | <0·0001 |
|  | AS-AQ | 2·24 (0·28-4·89) | *ref* | 89·5% (17/19) | *ref* | 2·01 (0·72-7·79) | *ref* | 94·7% (18/19) | *ref* |
|  | AS-AQ+PQ | 0 (0-0) | 0·832 | 20·0% (4/20) | <0·0001 | 0 (0-0) | 0·512 | 10% (2/20) | <0·0001 |
| Day 28 | AL | 0 (0-0) | *ref* | 5·0% (1/20) | *ref* | 0·21 (0·05-1·48) | *ref* | 80% (16/20) | *ref* |
|  | AL-AQ | 0 (0-0) | *nc* | 0% (0/18) | 0·526 | 0·21 (0-0·63) | 0·310 | 72·2% (13/18) | 0·427 |
|  | AL-AQ+PQ | 0 (0-0) | *nc* | 5·3% (1/19) | 0·744 | 0 (0-0) | *nc* | 0% (0/19) | <0·0001 |
|  | AS-AQ | 0·45 (0·12-3·21) | *ref* | 78·9% (15/19) | *ref* | 0·44 (0·13-4·21) | *ref* | 89·5% (17/19) | *ref* |
|  | AS-AQ+PQ | 0 (0-0) | 0·663 | 5·0% (1/20) | <0·0001 | 0 (0-0) | *nc* | 0% (0/20) | <0·0001 |

P-values are for differences between artemisinin-based combination therapy matched group comparison (i.e., artemether–lumefantrine vs artemether–lumefantrine-amodiaquine and artemether–lumefantrine-amodiaquine plus primaquine, artesunate-amodiaquine vs artesunate-amodiaquine plus primaquine). Density was compared using regression analyses of log10 transformed density values, with adjustment for baseline densities. Prevalence was compared with one sided Fishers exact tests. For the calculation of gametocyte prevalence, samples were classified as negative for a particular gametocyte sex if the estimated density of in gametocytes of that sex was less than 0·01 gametocytes per μL (i.e. one gametocyte per 100 μL of blood sample). · = not tested. *ref* = reference group, *nc* = not calculable. AL = artemether-lumefantrine; AL-AQ = artemether-lumefantrine-amodiaquine; AL-AQ+PQ = artemether-lumefantrine-amodiaquine plus primaquine; AS-AQ = artesunate-amodiaquine; AS-AQ+PQ = artesunate-amodiaquine plus primaquine

### Supplementary Figure 4. Proportion of gametocytes that were male

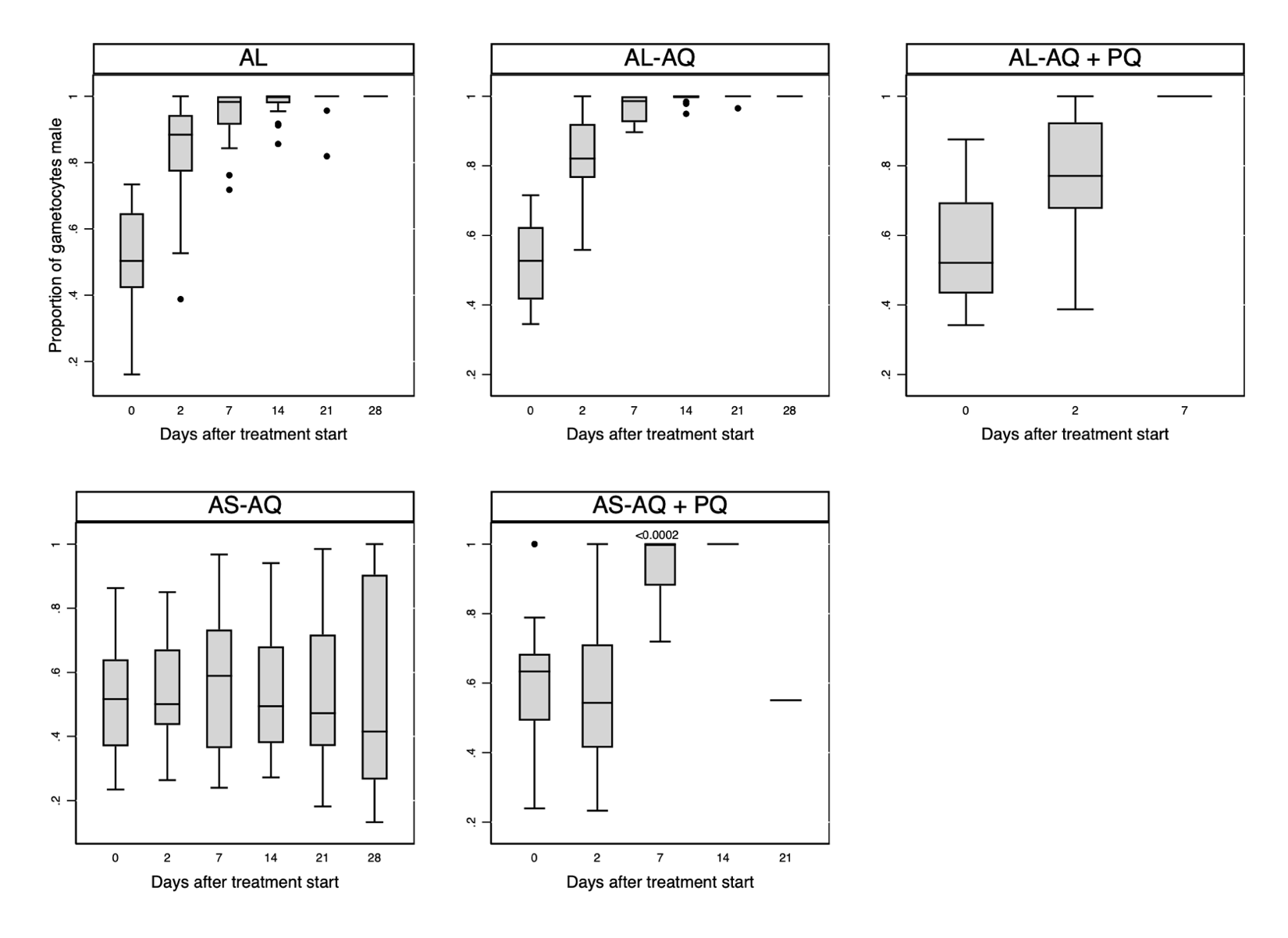

The proportion of gametocytes that were male was calculated for all values with total gametocyte densities of 0.2/µL and over, as described previously.(1) P-values (<0.05) for differences between treatment groups AL-AQ, AL-AQ+PQ and the AL reference group, and between AS-AQ+PQ and the AS-AQ reference group, were calculated using Wilcoxon rank sum tests. AL = artemether-lumefantrine; AL-AQ = artemether-lumefantrine-amodiaquine; AL-AQ+PQ = artemether-lumefantrine-amodiaquine plus primaquine; AS-AQ = artesunate-amodiaquine; AS-AQ+PQ = artesunate-amodiaquine plus primaquine.

### Supplementary Table 9. Gametocyte infectivity

| Day of follow-up | Treatment arm | Odds ratio (95% CI) | P-value |
| --- | --- | --- | --- |
| Day 0 | AL | 1 | *ref* |
|  | AL-AQ | 0·64 (0·47-0·89) | 0·008 |
|  | AL-AQ+PQ | 0·19 (0·13-0·28) | <0·0001 |
|  | AS-AQ | 1 | *ref* |
|  | AS-AQ+PQ | 1·71 (1·29-2·27) | <0·0001 |
| Day 2 | AL | 1 | *ref* |
|  | AL-AQ | 1·3 (0·15-11·27) | 0·808 |
|  | AL-AQ+PQ | *nc* | *nc* |
|  | AS-AQ | 1 | *ref* |
|  | AS-AQ+PQ | *nc* | *nc* |

Odds ratios are for the change in mosquito infection rate in the ALAQ and ALAQ+PQ groups compared to the reference (AL) group and the AS-AQ+PQ group compared to the reference (AS-AQ) group with adjustment for gametocyte densities. At later timepoints, there were too few infected mosquitoes to calculate the odd ratios. *nc* = not calculable, no observations (too few infected mosquitoes for convergence), *ref* = reference, AL = artemether-lumefantrine; AL-AQ = artemether-lumefantrine-amodiaquine; AL-AQ+PQ = artemether-lumefantrine-amodiaquine plus primaquine; AS-AQ = artesunate-amodiaquine; AS-AQ+PQ = artesunate-amodiaquine plus primaquine.

### Supplementary Table 10. Haemoglobin density

Haemoglobin density and percent reduction in haemoglobin density (relative to baseline) were compared within treatment arms (p-value¥) using paired t-tests (with day 0 as reference for percent change) and between treatment arms (p-value†) using linear regression (for density, adjusted for baseline Hb density) or two-way t-tests (for percent reduction). · = not tested. *ref* = reference group, AL = artemether-lumefantrine; AL-AQ = artemether-lumefantrine-amodiaquine; AL-AQ+PQ = artemether-lumefantrine-amodiaquine plus primaquine; AS-AQ = artesunate-amodiaquine; AS-AQ+PQ = artesunate- amodiaquine plus primaquine.

| Day of follow-up | Treatment arm | Mean g/dL (range) | p-value^¥^ | p-value^†^ | Percent change from day 0 | | | |
| --- | --- | --- | --- | --- | --- | --- | --- | --- |
|  |  |  |  |  | Mean (lower/upper 95% CI) | Range | p-value^¥^ | p-value^†^ |
| Day 0 | **Overall** | 12·0 (10·1-14·9) | · | · | · | · | · | · |
|  | **AL** | 12·5 (10·4-14·9) | *ref* | *ref* | · | · | · | · |
|  | **AL-AQ** | 12 (10·1-14·9) | *ref* | 0·283 | · | · | · | · |
|  | **AL-AQ+PQ** | 11·7 (10·4-13·4) | *ref* | 0·031 | · | · | · | · |
|  | **AS-AQ** | 11·8 (10·5-14·5) | *ref* | *ref* | · | · | · | · |
|  | **AS-AQ+PQ** | 11·9 (10·1-13·8) | *ref* | 0·748 | · | · | · | · |
| Day 1 | **AL** | 11·9 (9·5-15·5) | 0·0354 | *ref* | -4·41% (-8·28 / -0·54) | -25·17/8·65 | 0·0278 | *ref* |
|  | **AL-AQ** | 11·7 (10-15) | 0·1068 | 0·530 | -2·40% (-5·52 / 0·71) | -12·03/8·18 | 0·1232 | 0·4038 |
|  | **AL-AQ+PQ** | 11·1 (9·8-14) | 0·0005 | 0·761 | -5·36% (-7·93 / -2·79) | -15·97/4·48 | 0·0003 | 0·6719 |
|  | **AS-AQ** | 11·4 (9-15·5) | 0·0069 | *ref* | -3·79% (-6·42 / -1·17) | -14·29/7·62 | 0·0070 | *ref* |
|  | **AS-AQ+PQ** | 11·5 (9·6-13·6) | 0·0408 | 0·821 | -3·19% (-6·35 / -0·04) | -13·79/8·13 | 0·0474 | 0·7617 |
| Day 2 | **AL** | 12 (10·1-15) | 0·0006 | *ref* | -4·02% (-6·02 / -2·02) | -14·50/4·20 | 0·0005 | *ref* |
|  | **AL-AQ** | 11·5 (9·6-13·2) | 0·0007 | 0·411 | -4·83% (-7·25 / -2·41) | -12·71/4·55 | 0·0005 | 0·5890 |
|  | **AL-AQ+PQ** | 11 (9·6-12·5) | <0·00001 | 0·167 | -5·42% (-7·18 / -3·66) | -12·31/3·67 | <0·00001 | 0·2779 |
|  | **AS-AQ** | 11·2 (9·9-13·5) | <0·00001 | *ref* | -5·59% (-7·54 / -3·64) | -14·63/1·74 | <0·00001 | *ref* |
|  | **AS-AQ+PQ** | 11·4 (10·2-13·6) | 0·0005 | 0·288 | -4·36% (-6·50 / -2·23) | -13·33/4·27 | 0·0004 | 0·3797 |
| Day 7 | **AL** | 12·3 (10·1-15·9) | 0·1394 | *ref* | -1·52% (-3·66 / 0·62) | -11·40/6·71 | 0·1543 | *ref* |
|  | **AL-AQ** | 12 (10·4-14·2) | 0·5061 | 0·947 | -0·55% (-3·40 / 2·31) | -7·52/16·35 | 0·6917 | 0·5649 |
|  | **AL-AQ+PQ** | 11·6 (10·4-13·7) | 0·7899 | 0·629 | -0·23% (-2·30 / 1·84) | -12·61/5·50 | 0·8165 | 0·3715 |
|  | **AS-AQ** | 11·6 (9·7-14·2) | 0·1751 | *ref* | -1·53% (-4·05 / 0·99) | -13·01/8·33 | 0·2191 | *ref* |
|  | **AS-AQ+PQ** | 11·8 (10·2-13·3) | 0·1723 | 0·802 | -1·31% (-3·61 / 0·99) | -10·53/6·25 | 0·2478 | 0·8933 |
| Day 14 | **AL** | 12·6 (10·5-15·5) | 0·4351 | *ref* | 1·37% (-1·50 / 4·23) | -10·29/13·21 | 0·3309 | *ref* |
|  | **AL-AQ** | 12·3 (11·1-14·6) | 0·2260 | 0·787 | 1·86% (-0·85 / 4·57) | -8·46/11·54 | 0·1657 | 0·7948 |
|  | **AL-AQ+PQ** | 11·9 (10·9-13·2) | 0·1487 | 0·568 | 2·40% (-0·58 / 5·38) | -10·77/11·30 | 0·1082 | 0·6030 |
|  | **AS-AQ** | 11·9 (10·4-14·8) | 0·2327 | *ref* | 1·19% (-0·63 / 3·00) | -4·07/7·62 | 0·1869 | *ref* |
|  | **AS-AQ+PQ** | 12·5 (10·5-13·8) | 0·0026 | 0·021 | 4·64% (1·90 / 7·37) | -6·67/19·27 | 0·0021 | 0·0357 |
| Day 21 | **AL** | 12·6 (10·6-15·2) | 0·5576 | *ref* | 1·65% (-2·79 / 6·09) | -10·92/21·70 | 0·4462 | *ref* |
|  | **AL-AQ** | 12·6 (11-14·7) | 0·0208 | 0·481 | 4·60% (0·98 / 8·21) | -4·17/25·64 | 0·0157 | 0·2938 |
|  | **AL-AQ+PQ** | 12·1 (11·1-13·4) | 0·0092 | 0·943 | 4·34% (1·41 / 7·27) | -9·23/12·50 | 0·0062 | 0·3075 |
|  | **AS-AQ** | 12·2 (10·3-14·9) | 0·0429 | *ref* | 3·77% (0·48 / 7·07) | -14·39/18·10 | 0·0272 | *ref* |
|  | **AS-AQ+PQ** | 12·3 (10·3-13·6) | 0·0087 | 0·923 | 3·06% (0·89 / 5·23) | -2·68/11·93 | 0·0083 | 0·7026 |
| Day 28 | **AL** | 12·8 (11·3-15·2) | 0·0838 | *ref* | 2·75% (-0·05 / 5·55) | -7·69/16·04 | 0·0535 | *ref* |
|  | **AL-AQ** | 12·7 (10·6-14·5) | 0·0103 | 0·436 | 5·33% (1·50 / 9·17) | -9·40/18·18 | 0·0093 | 0·2540 |
|  | **AL-AQ+PQ** | 12·1 (11·3-13·1) | 0·0233 | 0·266 | 3·71% (0·62 / 6·80) | -5·04/21·15 | 0·0213 | 0·6328 |
|  | **AS-AQ** | 12·4 (11·1-14·9) | 0·0009 | *ref* | 5·67% (2·64 / 8·71) | -1·63/19·64 | 0·0010 | *ref* |
|  | **AS-AQ+PQ** | 12·5 (10·8-14·3) | 0·0029 | 0·850 | 4·86% (1·83 / 7·88) | -4·31/15·60 | 0·0033 | 0·6914 |

Supplementary Figure 5. Absolute haemoglobin density
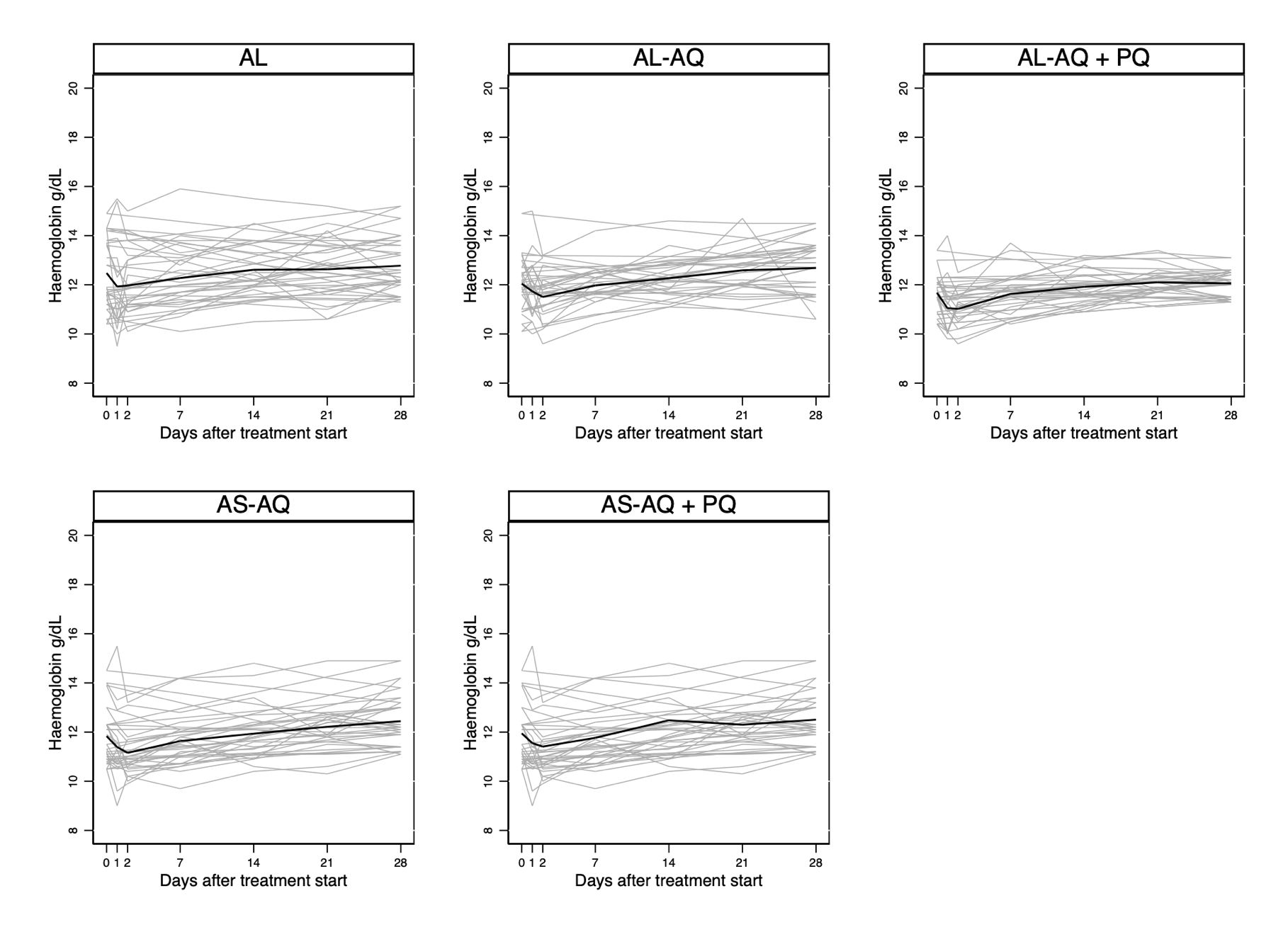

Absolute haemoglobin density is given in grams per dL (y-axis, from 8-20 g/dL) and is indicated for each participant individually with grey lines. The single black line shows the mean absolute haemoglobin density. P-values are presented in Supplementary table 8. AL = artemether-lumefantrine; AL-AQ = artemether-lumefantrine-amodiaquine; AL-AQ+PQ = artemether-lumefantrine-amodiaquine plus primaquine; AS-AQ = artesunate-amodiaquine; AS-AQ+PQ = artesunate-amodiaquine plus primaquine.

### Supplementary Table 11. Biochemistry

| Day of follow-up | Treatment arm | Mean ALT U/L (range) | p-value¥ | p-value† | Mean AST U/L (range) | p-value¥ | p-value† | Mean creatine mg/dL (range) | p-value¥ | p-value† |
| --- | --- | --- | --- | --- | --- | --- | --- | --- | --- | --- |
| Day 0 | **Overall** | 23·45 (8-157) | · | · | 29·24 (4-215) | · | · | 0·64 (0·2-1·2) | · | · |
|  | **AL** | 20·90 (12-37) | *ref* | *ref* | 23·95 (4-40) | *ref* | *ref* | 0·63 (0·2-1·2) | *ref* | *ref* |
|  | **AL-AQ** | 19·15 (10-27) | *ref* | 0·387 | 25·45 (13-35) | *ref* | 0·562 | 0·77 (0·3-1·2) | *ref* | 0·055 |
|  | **AL-AQ+PQ** | 21·50 (8-67) | *ref* | 0·881 | 30·30 (16-62) | *ref* | 0·065 | 0·66 (0·2-1·2) | *ref* | 0·650 |
|  | **AS-AQ** | 25·50 (7-117) | *ref* | *ref* | 27·25 (2-63) | *ref* | *ref* | 0·58 (0·2-1·0) | *ref* | *ref* |
|  | **AS-AQ+PQ** | 24·75 (5-118) | *ref* | 0·921 | 26·75 (6-42) | *ref* | 0·881 | 0·66 (0·2-1·2) | *ref* | 0·303 |
| Day 2 | **AL** | 18·35 (9-40) | 0·0694 | *ref* | 21·80 (4-40) | 0·4851 | *ref* | 0·77 (0·1-1·4) | 0·0280 | *ref* |
|  | **AL-AQ** | 22·00 (8-57) | 0·2813 | 0·125 | 28·37 (5-48) | 0·1114 | 0·045 | 0·77 (0·2-1·3) | 0·9404 | 0·279 |
|  | **AL-AQ+PQ** | 29·16 (7-164) | 0·2216 | 0·099 | 31·32 (11-94) | 0·8206 | 0·226 | 0·61 (0·3-1·0) | 0·3205 | 0·017 |
|  | **AS-AQ** | 30·10 (12-126) | 0·5847 | *ref* | 34·80 (8-163) | 0·3167 | *ref* | 0·64 (0·3-1·0) | 0·3306 | *ref* |
|  | **AS-AQ+PQ** | 28·10 (7-105) | 0·1422 | 0·821 | 28·90 (11-52) | 0·5094 | 0·484 | 0·63 (0·2-1·2) | 0·6319 | 0·573 |
| Day 7 | **AL** | 20·40 (11-33) | 0·7623 | *ref* | 24·55 (10-37) | 0·8239 | *ref* | 0·61 (0·3-1·0) | 0·6154 | *ref* |
|  | **AL-AQ** | 22·00 (7-44) | 0·1483 | 0·276 | 28·94 (8-44) | 0·1364 | 0·143 | 0·74 (0·4-1·2) | 0·6901 | 0·259 |
|  | **AL-AQ+PQ** | 26·05 (8-129) | 0·4681 | 0·380 | 29·84 (8-110) | 0·8915 | 0·628 | 0·62 (0·4-0·8) | 0·4581 | 0·915 |
|  | **AS-AQ** | 26·45 (9-92) | 0·8855 | *ref* | 27·30 (10-38) | 0·9835 | *ref* | 0·65 (0·2-1·0) | 0·2183 | *ref* |
|  | **AS-AQ+PQ** | 25·20 (13-57) | 0·9068 | 0·810 | 29·75 (12-49) | 0·1975 | 0·293 | 0·72 (0·2-1·0) | 0·1907 | 0·476 |
| Day 14 | **AL** | 19·45 (10-38) | 0·2991 | *ref* | 24·95 (9-39) | 0·7192 | *ref* | 0·64 (0·2-1·0) | 0·8152 | *ref* |
|  | **AL-AQ** | 28·72 (10-157) | 0·2054 | 0·137 | 36·17 (19-215) | 0·2846 | 0·300 | 0·70 (0·2-1·0) | 0·2615 | 0·861 |
|  | **AL-AQ+PQ** | 25·11 (8-112) | 0·6021 | 0·354 | 33·95 (15-86) | 0·4069 | 0·140 | 0·60 (0·3-0·8) | 0·3105 | 0·407 |
|  | **AS-AQ** | 25·47 (15-65) | 0·9046 | *ref* | 26·89 (4-42) | 0·9360 | *ref* | 0·62 (0·2-1·2) | 0·3620 | *ref* |
|  | **AS-AQ+PQ** | 19·20 (9-30) | 0·2683 | 0·043 | 25·05 (5-35) | 0·3725 | 0·520 | 0·64 (0·4-1·2) | 0·7263 | 0·742 |

Alanine aminotransferase (ALT), aspartate aminotransferase (AST) and creatinine were compared within treatment arms (p-value[^§^](https://www.thelancet.com/journals/lanmic/article/PIIS2666-5247(21)00356-6/fulltext)) using paired t-tests (with day 0 as reference) and between treatment arms (p-value[^¶^](https://www.thelancet.com/journals/lanmic/article/PIIS2666-5247(21)00356-6/fulltext)) using linear regression (adjusted for baseline levels). *Ref* = reference, · = not tested. AL = artemether-lumefantrine; AL-AQ = artemether-lumefantrine-amodiaquine; AL-AQ+PQ = artemether-lumefantrine-amodiaquine plus primaquine; AS-AQ = artesunate-amodiaquine; AS-AQ+PQ = artesunate-amodiaquine plus primaquine.

### Supplementary Table 12. All adverse events

| **Description** | **Total (n=100)** | **AL (n=20)** | **AL-AQ (n=20)** | **AL-AQ+PQ (n=20)** | **AS-AQ (n=20)** | **AS-AQ+PQ (n=20)** |
| --- | --- | --- | --- | --- | --- | --- |
| Abdominal pain | 23^21^(5) | 3^3^ | 5^5^(1) | 4^4^(1) | 5^5^(3) | 6^4^ |
| Acute respiratory infection | 26^1^(17) | 6(2) | 2(2) | 7(4) | 6^1^(4) | 5(5) |
| Allergic contact dermatitis | 1(1) | 0 | 0 | 1(1) | 0 | 0 |
| Anemia | 1^1^ | 0 | 1^1^ | 0 | 0 | 0 |
| Asthenia | 16^16^(5) | 2^2^ | 4^4^(2) | 3^3^ | 4^4^(2) | 3^3^(1) |
| Chills | 2^2^ | 1^1^ | 0 | 1^1^ | 0 | 0 |
| Conjunctivitis | 3(3) | 1(1) | 1(1) | 1(1) | 0 | 0 |
| Cough | 9^2^(4) | 2^1^(1) | 1(1) | 1 | 4^1^(1) | 1(1) |
| Diarrhea | 4^4^(1) | 0 | 2^2^(1) | 0 | 1^1^ | 1^1^ |
| Drowsiness | 5^5^(1) | 1^1^ | 1^1^ | 2^2^(1) | 1^1^ | 0 |
| Dyspnea | 1^1^ | 0 | 0 | 1^1^ | 0 | 0 |
| Eczema | 1(1) | 0 | 1(1) | 0 | 0 | 0 |
| Elevation of ALT/ GPT | 5^2^(1) | 0 | 1 | 2^1^(1) | 2^1^ | 0 |
| Elevation of ASAT/GOT | 5^1^ | 0 | 1 | 3^1^ | 1 | 0 |
| Elevation of creatinemia | 3^2^ | 2^1^ | 0 | 0 | 0 | 1^1^ |
| Fatigue | 4^3^ | 2^1^ | 0 | 2^2^ | 0 | 0 |
| Fever | 2^1^(1) | 1^1^(1) | 0 | 0 | 0 | 1 |
| Food indigestion | 1(1) | 0 | 0 | 0 | 1(1) | 0 |
| Headaches | 51^38^(23) | 8^7^(3) | 10^8^(5) | 12^10^(7) | 8^6^(4) | 13^7^(4) |
| Hyperleukocytosis | 4^1^ | 1 | 0 | 2^1^ | 0 | 1 |
| Leucopenia | 3^3^ | 0 | 0 | 0 | 2^2^ | 1^1^ |
| Liquid diarrhea | 1^1^(1) | 0 | 1^1^(1) | 0 | 0 | 0 |
| Localized left arm pain | 1 | 0 | 0 | 1 | 0 | 0 |
| Loss of appetite | 10^10^ | 0 | 3^3^ | 2^2^ | 5^5^ | 0 |
| Low back pain | 1 | 0 | 1 | 0 | 0 | 0 |
| Muscular pain | 10^8^(1) | 1(1) | 1^1^ | 3^3^ | 3^2^ | 2^2^ |
| Nausea | 17^17^(2) | 3^3^ | 3^3^(1) | 3^3^ | 6^6^(1) | 2^2^ |
| Rhinitis | 3(1) | 1(1) | 2 | 0 | 0 | 0 |
| Rhinorrhea | 13(1) | 3 | 3 | 2 | 3 | 2(1) |
| Traumatic wound right food | 1(1) | 0 | 1(1) | 0 | 0 | 0 |
| Uncomplicated malaria | 1(1) | 1(1) | 0 | 0 | 0 | 0 |
| Vertigo | 19^18^(6) | 2^1^ | 3^3^(1) | 6^6^(4) | 4^4^(1) | 4^4^ |
| Vomiting | 15^14^(3) | 4^4^(1) | 4^3^(1) | 3^3^ | 3^3^(1) | 1^1^ |
| **ALL** | *262^172^* | *45^26^* | *52^35^* | *62^43^* | *59^42^* | *44^26^* |
| ***MILD*** | *181^129^* | *33^21^* | *33^22^* | *42^32^* | *41^30^* | *32^24^* |
| ***MODERATE*** | *81^43^* | *12^5^* | *19^13^* | *20^11^* | *18^12^* | *12^2^* |

85/100 participants experienced a total of 262 adverse events over the course of the trial; 181 categorised for severity by the study clinician (in accordance with the study protocol and data safety and monitoring charter) as ‘mild’ and 81 as ‘moderate’. No severe adverse events or serious adverse events (SAE) occurred during the trial. The frequency of all AEs is given outside parentheses, with the frequency of moderate AEs in parentheses. The frequency of AEs that were related to drug treatment (defined as probably, possibly or definitely related to treatment) is given in superscript. 172 of the 262 AEs were classified as possibly, probably or definitely related to the study drug; of these, 129/169 were mild and 43/169 were moderate.
