## Supplementary material for "Artemether-lumefantrine-amodiaquine or artesunate-amodiaquine combined with single low-dose primaquine to reduce *Plasmodium falciparum* malaria transmission in Ouélessébougou, Mali: a five-arm, phase 2, single-blind, randomised clinical trial": Study protocol

**Principal Investigators**

| Prof Alassane Dicko  Faculty of Pharmacy and Faculty of Medicine and Dentistry, University of Sciences Techniques and Technologies of Bamako, Mali  Prof. Dr. Chris Drakeley  London School of Hygiene & Tropical Medicine, United Kingdom |  |
| --- | --- |
| **Co-investigators: Mali**  Dr. Halimatou Diawara  Dr. Almahamoudou Mahamar  Dr. Harouna Soumare  Dr. Djibrilla Issiaka  Prof. Cheick Traore  Malaria Research and Training Centre, Faculty of Pharmacy and Faculty of Medicine and Dentistry, University of Sciences Techniques and Technologies of Bamako, Mali | **Co-investigators: International**  Prof. Teun Bousema  Drs. Merel Smit  Radboud University Medical Center, Nijmegen, The Netherlands  Dr. William Stone  Dr. Leen Vanheer  London School of Hygiene & Tropical Medicine, United Kingdom |

London School of Hygiene & Tropical Medicine is the main research sponsor for this study. For further information regarding the sponsorship conditions, please contact the Research Governance and Integrity Office:

London School of Hygiene & Tropical Medicine

Keppel Street

London WC1E 7HT

*Planned start date:	September 2022*

*Planned end date: July 2023*

9.1.7.	Confirmatory pregnancy test at the end of follow-up	18

### 1. LIST OF ABBREVIATIONS

ACT	Artemisinin-based combination therapy

AE	Adverse event

AL	Artemether-Lumefantrine

AQ	Amodiaquine

AS	Artesunate

ASAQ	Artesunate-Amodiaquine

AUC	Area under the curve

CBC	Complete Blood Count

CRF	Case report form

DNA	Deoxyribonucleic acid

DP	Dihydroartemisinin-Piperaquine

DSMB	Data and Safety Monitoring Board

EC	Ethics Committee

EDTA	Ethylenediaminetetraacetic acid

G6PD	Glucose-6-phosphatase dehydrogenase

GCP	Good clinical practice

Hb	Haemoglobin

Hct	Haematocrit

LSHTM	London School of Hygiene and Tropical Medicine

MACS	Magnetic-activated cell sorting

MFA	Membrane feeding assay

MRTC	Malaria Research and Training Centre (Bamako, Mali)

*P. falciparum	Plasmodium falciparum*

PA	Pyronaridine-Artesunate

PCR	Polymerase chain reaction

PQ	Primaquine

RBCs	Red blood cells

RT-qPCR	Quantitative reverse transcriptase polymerase chain reaction

SAE	Serious Adverse Event

SLD	Single low dose

SOP	Standard operating procedure

SUSAR	Serious and Unexpected Suspected Adverse Reaction

TACT	Triple Artemisinin-based Combination Therapy

UP	Unanticipated Problem

UPnonAE	Unanticipated Problem that is not an Adverse Event

WHO	World Health Organisation

### 2. STUDY SYNOPSIS

Artemisinin Combination Therapies (ACT) are the first-line treatment for uncomplicated *Plasmodium falciparum* malaria. ACTs rapidly clear asexual *P. falciparum* parasites, responsible for clinical symptoms, but have limited activity against mature gametocytes, which are the only life stages capable of transmitting to mosquitoes. To reduce *P. falciparum* transmission, the World Health Organisation (WHO) recommends the addition of a single low dose of primaquine (0.25 mg/kg), a potent and fast-acting gametocytocidal drug, to ACT [1]. Although Artesunate-Amodiaquine (ASAQ) is a commonly used ACT, limited available data suggest poor activity against gametocytes [2] and the added benefit of SLD PQ in combination with ASAQ remains unknown. We previously showed that Artemether-Lumefantrine (AL) is a more potent ACT in terms of gametocyte clearance and transmission reduction efficacy compared to Dihydroartemisinin-Piperaquine (DP) [3], [4] [NECTAR3, in preparation]. Recently, triple artemisinin-based combination therapies (TACT) such as Artemether-Lumefantrine plus Amodiaquine (ALAQ) have been proposed to delay the emergence of drug resistance and to provide efficacious treatment for multi-drug resistance *P. falciparum* infections [5], [6]. The transmission reducing efficacy of ALAQ or ALAQ-PQ has not yet been tested.

In this study we aim to determine transmission reducing efficacy of Artesunate-Amodiaquine (ASAQ) with and without a single low dose (0.25mg/kg) of Primaquine (SLD PQ) and Artemether-Lumefantrine (AL) with and without Amodiaquine (AQ) and Primaquine (PQ). To achieve this, we propose to conduct a five-arm, single-blind, randomised control trial to test the gametocytocidal and transmission-blocking efficacy of SLD PQ in combination with Artesunate-Amodiaquine and Artemether-Lumefantrine-Amodiaquine. Individuals will be recruited to an AL-only arm as an internal control for the ALAQ arm, and for comparison to recent trials at the same site [NECTAR3, in preparation]. Individuals who meet inclusion criteria will be randomised in a 1:1:1:1:1 ratio to: ASAQ, ASAQ-PQ, AL, ALAQ, ALAQ-PQ.

For participation in the trial, individuals between 10-50 years will be recruited from villages around Ouelessebougou, near Bamako in Mali. Written, informed consent will be obtained. Inclusion criteria include presence of ≥16 gametocytes/µL, absence of other non-*P. falciparum* species on blood film, absence of symptomatic falciparum malaria, haemoglobin ≥ 10 g/dL, and no evidence of acute severe or chronic disease or pregnancy. Recruitment will continue until 100 individuals who meet the inclusion criteria are recruited and randomised. G6PD enzyme status will not be tested.

Participants will be followed for 28 days. Blood samples will be taken pre-treatment, and at days 2, 7, 14, 21 and 28 post-treatment for parasitology, genetic, haematology and biochemistry analysis. MFA will be conducted pre-treatment, and at days 2, 7 and 14 post-treatment. MFA will also be conducted at day 21 for any individuals infectious at day 7 or 14; and at day 28 for any individuals infectious at day 14 or 21. Magnetic-activated cell sorting (MACS) to enrich for gametocytes will be done at two timepoints per individual to determine per-gametocyte infectivity and confirm whether PQ sterilizes gametocytes, followed by MFA. The primary outcome will be the reduction in mosquito infection rate within each arm at day 2 post-treatment. Secondary outcomes include within and between arm comparisons in other mosquito infection, gametocyte, safety, and biochemical parameters.

### 3. STUDY BACKGROUND AND RATIONALE

#### 3.1. Background

Worldwide malaria morbidity and mortality remains unacceptably high, with an estimated 241 million cases in 2020, a substantial increase from 214 million in 2015 [7]. In addition to this, the emergence and spread of resistance to first-line drugs is threatening to further increase this rise in malaria cases and deaths. Therefore, there is a clear need for interventions that reduce malaria transmission.

Artemisinin combination-based therapies (ACTs) are the first-line treatment for uncomplicated *P. falciparum* malaria; however, they have very limited activity against mature gametocytes, the stages responsible for transmission to the mosquito. The 8-aminoquinolone primaquine is a potent and fast-acting gametocytocidal drug and is currently the only drug recommended for blocking malaria transmission. To reduce *P. falciparum* transmission, the World Health Organisation (WHO) recommends the addition of a single low dose of primaquine (SLD PQ) (0.25 mg/kg), to ACT. The benefit of adding SLD PQ has been assessed with Dihydroartemisinin-Piperaquine (DP), Pyronaridine-Artesunate (PA) and Artemether-Lumefantrine (AL), concluding effective blockage of transmission within 48 hours [3], [8] [NECTAR3, in preparation]. However, the added benefit of combining PQ with Artesunate-Amodiaquine (ASAQ) has not yet been tested, despite ASAQ being the first-line ACT in many countries [7]. Importantly, an individual-level meta-analysis of gametocyte data before and after ACT treatment suggested that ASAQ may be inefficient in clearing gametocytes with a fourfold higher odds of gametocyte carriage compared to the most widely used ACT that is AL [2].

The stagnation of progress toward reducing global malaria morbidity and mortality may be compounded by the emergence of artemisinin resistance. Artemisinin resistance, characterized by slow parasite clearance [9], [10] because of reduced susceptibility of ring-stage parasites [11], first emerged in Southeast Asia, a region that has been the epicentre for resistance development against all the major anti-malaria drugs in the past. In many areas with artemisinin resistance, resistance to the partner drug has now emerged as well, leading to a rapid rise in treatment failures following ACT [12], [13].

Artemisinin resistance is mediated by mutations in the propeller region of the *Kelch 13* (K13) gene. Recently, K13 mutations and associated delayed clearance of parasites have been found in East-Africa as well [14], [15]. Wider spread of artemisinin resistance across the African continent, which bears the vast majority of the global malaria burden, could be disastrous. In order to avert a public health emergency, there is an urgent need for alternative treatments with combinations of existing drugs that could be deployed rapidly. Triple Artemisinin-based Combination Therapies (TACT), which combine an existing ACT with a second partner drug that is slowly eliminated, could be part of such an approach.

The rationale for TACT is similar to the one underlying ACT. By combining drugs with different treatment targets and/or resistance mechanisms, the probability that resistance will emerge to the combination is very low (i.e., the product of the probabilities for each individual drug). TACT could counteract resistance emergence and provide efficacious treatment against drug resistant infections. Artemether-Lumefantrine with Amodiaquine (ALAQ) is one such TACT and has been proven a safe, well-tolerated and efficacious treatment for uncomplicated *P. falciparum* malaria, including in areas with artemisinin and partner-drug resistance [5], [6]. The effect of ALAQ on mature gametocytes and infectivity is unknown, as well as the added benefit of combining PQ with ALAQ. Given that a major rationale for using TACT is a reduction in the transmission of artemisinin resistance, it is pressing to understand the impact of TACT on gametocyte production and infectivity.

#### 3.2. Rationale

Artesunate-Amodiaquine (ASAQ) is a commonly used ACT, however, its transmission reducing efficacy remains largely unknown. Studies microscopically assessing gametocyte carriage post-ASAQ treatment reached very different conclusions, ranging from increased gametocyte carriage until day 3 [16] or day 7 [17] post-treatment to an immediate decrease in gametocytaemia [18]. Compared to AL, ASAQ appears to be less efficient in preventing post-treatment gametocyte carriage [2]. In addition, it is important to note that gametocyte carriage is a poor measurement for human-to-mosquito transmission, as antimalarial drugs can sterilise gametocytes before removing them from the blood stream [19] and transmissible gametocytes can persist at submicroscopic densities [20]. Membrane feeding assays (MFA) are therefore the most reliable method to assess transmission. MFA can be supported by magnetic enrichment of gametocytes that can allow for assessments of infectivity of increased gametocyte densities [21] and differentiate between lack of infectivity because of gametocyte densities that are too low to permit transmission and gametocyte sterilization [22].This is relevant since uncertainties remain about the exact working mechanism of transmission-blocking antimalarial drugs that may clear gametocytes, distort their sex-ratio or sterilize gametocytes without clearance from circulation [19]. Understanding the mode by which transmission is prevented can support drug development.

Only one study has performed MFA after ASAQ treatment, reporting an increase in transmission [17]. If there is indeed substantial transmission after treatment with ASAQ, the addition of a single low dose PQ may be considered. To date, the added benefit of PQ when used in combination with ASAQ is unknown.

We previously showed that Artemether-Lumefantrine (AL) is a more potent ACT in terms of gametocyte clearance and transmission reduction efficacy compared to Dihydroartemisinin-Piperaquine (DP) [3], [4], [NECTAR3, in preparation]. It is currently unclear what the reason is for this marked difference and whether gametocyte-sterilization plays a role or the effect is driven by gametocyte-clearance. Recently, triple artemisinin-based combination therapies (TACT) such as Artemether-Lumefantrine plus Amodiaquine (ALAQ), which combine an existing ACT with a second partner drug that is slowly eliminated, have been proposed to provide efficacious treatment for multidrug-resistant *P. falciparum* infections and to delay the emergence of drug resistance [5], [6]. The transmission reducing efficacy of ALAQ or ALAQ combined with a single dose of PQ has not yet been tested.

Therefore, we propose to conduct a five-arm, phase 2, single-blind, randomised control trial to test the gametocytocidal and transmission-blocking efficacy of SLD PQ in combination with Artesunate-Amodiaquine and Artemether-Lumefantrine-Amodiaquine. Individuals will be recruited to an AL only arm as an internal control for the ALAQ arm, and for comparison to recent trial [NECTAR3, in prep]. Individuals who meet inclusion criteria will be randomised in a 1:1:1:1:1 ratio to: ASAQ, ASAQ-PQ, AL, ALAQ, ALAQ-PQ.

### 4. OBJECTIVES

#### 4.1. Primary specific objective

To assess the within-person reduction of infectivity of gametocytes following administration of ASAQ alone or with single-dose PQ and AL alone or with AQ and ALAQ alone or with single-dose PQ in children and adults at day 2 post-treatment compared to pre-treatment (day 0).

#### 4.2. Secondary specific objectives

- To assess differences in other mosquito infectivity parameters (mosquito infection rate, change in infection rate, infectivity to mosquitoes, and oocyst density) following treatment with ASAQ, ASAQ-PQ, AL, ALAQ and ALAQ-PQ in children and adults at all feeding timepoints compared to pre-treatment (day 0) and between treatment matched arms (ASAQ vs ASAQ-PQ, AL vs ALAQ, ALAQ vs ALAQ-PQ).
- To assess differences in gametocyte parameters (prevalence, density, circulation time, area-under-the-curve, sex-ratio) following treatment with ASAQ, ASAQ-PQ, AL, ALAQ and ALAQ-PQ in children and adults at all feeding timepoints compared to pre-treatment (day 0) and between treatment matched arms (ASAQ vs ASAQ-PQ, AL vs ALAQ, ALAQ vs ALAQ-PQ).
- To assess differences in safety parameters (AE frequency, Hb density, median drop in Hb) following treatment with ASAQ, ASAQ-PQ, AL, ALAQ and ALAQ-PQ in children and adults at all feeding timepoints compared to pre-treatment (day 0) and between treatment matched arms (ASAQ vs ASAQ-PQ, AL vs ALAQ, ALAQ vs ALAQ-PQ).
- To assess differences in biochemical parameters (ALT, CBC, Creatinine) following treatment with ASAQ, ASAQ-PQ, AL, ALAQ and ALAQ-PQ in children and adults at all feeding timepoints compared to pre-treatment (day 0) and between treatment matched arms (ASAQ vs ASAQ-PQ, AL vs ALAQ, ALAQ vs ALAQ-PQ).

#### 4.3. Exploratory objectives

- To assess parasite genomic and transcriptomic variation at baseline and at select post-treatment timepoints.
- To assess human genomic variation (i.e., HBB type) and association with parasite measures.
- To assess the association of parasite and plasma biomarkers on malaria transmission efficiency.
- To assess the efficiency of *Plasmodium* transmission to mosquitoes before and after enrichment of gametocytes by MACS.
- To determine the association between gametocyte density and mosquito infection rates, before and after MACS enrichment, to estimate possible loss in per-gametocyte infectivity following treatment.

### 5. STUDY DESIGN

This is a five arm, single-blinded, randomized clinical trial. The study will be conducted according to the following steps for each individual:

Individuals aged 10-50 years will be invited to participate. They will be asked to provide informed consent and assent (for those under 18 years of age). After obtaining the informed consent and assent (if applicable), participants will undergo a screening including clinical evaluation and for the presence of microscopically detectable *P. falciparum* infection. Individuals with ≥16 gametocytes/µL will be recruited and go through additional screening including haemoglobin measurement, CBC, assessment of renal and liver functions, and pregnancy tests. Recruitment will continue until 100 infected individuals with microscopically detectable *P. falciparum* gametocytes (≥16/µL) are recruited and randomised.

Individuals who meet the inclusion and exclusion criteria will be randomised in a 1:1:1:1:1 ratio into five groups (arms); ASAQ, ASAQ-PQ (0.25mg/kg), AL, ALAQ, ALAQ-PQ (**Table 1**). Randomisation will be performed by the study pharmacist. All other staff will be blinded to the treatment allocation.

**Table 1. Treatment arms**

| **Arm** | **ACT** | **PQ (mg/kg)** | **Sample size** |
| --- | --- | --- | --- |
| 1 | ASAQ |  | 20 |
| 2 | ASAQ | 0.25 | 20 |
| 3 | AL |  | 20 |
| 4 | ALAQ |  | 20 |
| 5 | ALAQ | 0.25 | 20 |

Each participant will be followed for 28 days. Participants will receive a full clinical and parasitological examination on days 2, 7, 14, 21, 28 after receiving the first dose of the study drugs. Blood samples will be taken at all visits for parasitology (microscopy, and whole blood for qRT-PCR) and serology with additional samples for haematology and biochemistry analysis at days 0, 2 and 14. A clinical and haematological examination will also be performed on day 1, at which point no other samples will be collected.

Infectivity to locally reared mosquitoes will be assessed with membrane feeding assays at baseline, on the final day of treatment (day 2) and at days 7 and 14 for all participants. Feeds will then be performed at day 21 for any individuals infectious at day 7 or 14; and at day 28 for any individuals infectious at day 14 or 21. Magnetic-activated cell sorting (MACS) to enrich for gametocytes will be done at two timepoints per individual, followed by MFA.

In addition to this passive monitoring, participants will be prompted to contact the study coordinators if they feel sick at any point during follow up. A full clinical examination will be performed, and treatment will be provided according to national treatment guidelines.

#### 5.1. Study site

The Malaria Research and Training Centre (MRTC) in Bamako, Mali will use the field site of Ouelessebougou for recruitment. The MRTC has the unique position of being one of very few sites in Africa that can study malaria transmission endpoints. Since 2013 the MRTC successfully carried out similar trials at this site with great success, in collaboration with the University of California San Francisco, the London School of Hygiene and Tropical Medicine and the Radboud University Medical Center, Nijmegen, the Netherlands. In the 2013-2014 study, the MRTC demonstrated the efficacy of SLD PQ of doses of 0.125mg/kg or greater in combination with dihydroartemisinin-piperaquine for blocking P. falciparum transmission safely in G6PD normal Malian males from the age of 10-50 years [8]. MRTC has already established all the assays required for the study described in this protocol.

The MRTC is experienced in conducting Good Clinical Practice (GCP) compliant clinical trials including phase 1, 2, 3 and 4 of malaria vaccine and drug studies. Ouelessebougou village has been conducting ongoing clinical research since 2008 and is endemic for malaria with marked seasonality and a high burden of malaria in both children and adult populations. In recent years the prevalence of *P. falciparum* malaria in children under 5 years of age has ranged between 14% and 54% during the transmission season. The frequency of G6PD deficiency is in the range of 10 to 15%.

#### 5.2. Study population

The study population will be derived from individuals aged 10-50 years with asymptomatic *P. falciparum* malaria infection who agree to be screened for malaria infection.

For the randomised trial, we will aim to recruit 100 individuals. Eligible individuals will have microscopically detectable *P. falciparum* gametocyte densities of ≥16 gametocytes/µL.

#### 5.3. Intervention

Individuals who meet the inclusion criteria in the current study will be randomised in a 1:1:1:1:1 ratio into five groups (arms); ASAQ, ASAQ-PQ (0.25 mg/kg), AL, ALAQ, ALAQ-PQ (0.25 mg/kg). Randomisation will be performed by the study pharmacist. All staff other than the study pharmacist and clinician will be blinded to the treatment allocation. Participants will be told prior to enrolment that they can ask at any time which treatment arm they are in. The clinician or pharmacist will discuss this with them.

#### 5.4. Sample size calculation

For sample size considerations, we estimated the infectivity for participants in previous trials in the same study setting [3], [4], [8], [23] using a mixed effects logistic regression model that accounted for correlation between mosquito observations from the same participant. Based on a mixed effects logistic regression we looked at 100 simulations with an expected reduction in infectivity of 90%, an expected baseline proportion of infectivity of 17% for the average participant, and an expected intra-cluster correlation of 0.5. We estimated 88% empirical power to detect >80% reduction in infectivity with a one-tailed test with a 0.025 level of significance when including 20 participants and dissecting 50 mosquitos per participant at each timepoint.

#### 5.5. Inclusion criteria

- Age ≥ 10 years and ≤ 50 years
- Absence of symptomatic falciparum malaria, defined by fever on enrolment
- Presence of *P. falciparum* gametocytes on thick blood film at a density >16 gametocytes/µL (i.e. ≥ gametocytes recorded in the thick film against 500 white blood cells)
- Absence of other non-*P. falciparum* species on blood film
- Haemoglobin ≥ 10 g/dL
- Individuals weighing < = 80 kg
- No evidence of acute severe or chronic disease
- Written, informed consent

#### 5.6. Exclusion criteria

- Women who are pregnant or lactating (tested at baseline). Urine and/or serum pregnancy testing (β-hCG) will be used.
- Detection of a non-*P. falciparum* species by microscopy
- Previous reaction to study drugs / known allergy to study drugs, such as sudden high fevers, shaking or severe sore throat or ulcers in the mouth during treatment with Amodiaquine
- Current eye disease with retinal damage
- Signs of severe malaria, including hyperparasitaemia (defined as asexual parasitaemia > 100,000 parasites / µL)
- Signs of acute or chronic illness, including hepatitis
- The use of other medication (except for paracetamol and/or aspirin), including antacids, other medicines used to treat malaria, abnormal heart rhythm, depression or mental illness or HIV/AIDS, and medicines that have antibiotic/antifungal properties
- Use of antimalarial drugs over the past 7 days (as reported by the participant)
- Clinically significant illness (intercurrent illness e.g., pneumonia, pre-existing condition e.g., renal disease or HIV/AIDS, malignancy or conditions that may affect absorption of study medication e.g., severe diarrhoea or any signs of malnutrition as defined clinically)
- Signs of hepatic injury (such as nausea and/or abdominal pain associated with jaundice) or known severe liver disease (i.e., decompensated cirrhosis, Child Pugh stage B or C)
- Signs, symptoms or known renal impairment
- Clinically significant abnormal laboratory values as determined by history, physical examination, or routine blood chemistries and haematology values (laboratory guideline values for exclusion are haemoglobin < 10 g/dL, platelets < 50,000/μl, White Blood Cell count (WBC) < 2000/μl, serum creatinine >2.0mg/dL, or ALT more than 3 times the upper limit of normal for age.
- Blood transfusion in the last 90 days.
- Known Electrocardiogram (ECG) corrected QT interval of more than 450 ms
- Documented or self-reported history of cardiac conduction problems
- Documented or self-reported history of epileptic seizures

### 6. STUDY DRUGS

#### 6.1. Artesunate-Amodiaquine (ASAQ)

Participants in the ASAQ arm will receive a fixed-dose combination in tablets containing 50mg/135 mg or 100mg/270 mg of artesunate/amodiaquine (Fosun Pharma, Shanghai, China). Tablets will be administered according to manufacturer guidelines (**Table 2**).

**Table 2.** ASAQ dosing

| **Weight** | **Tablets** | **D0** | **D1** | **D2** |
| --- | --- | --- | --- | --- |
| 9 to < 18 kg | 50 mg AS/135 mg AQ base | 1 tab | 1 tab | 1 tab |
| 18 to < 36 kg | 100 mg AS/270 mg AQ base | 1 tab | 1 tab | 1 tab |
|  | blister pack of 3 tab |  |  |  |
| ≥ 36 kg | 100 mg AS/270 mg AQ base | 2 tab | 2 tab | 2 tab |
|  | blister pack of 6 tab |  |  |  |

#### 6.2. Primaquine (PQ)

Participants in the ASAQ-PQ arm will receive PQ (ACE Pharmaceuticals, Zeewolde, The Netherlands) at a single low dose of 0.25mg/kg as is currently recommended by the World Health Organization. The single dose of PQ will be given on day 0 together with ASAQ, administered in an aqueous solution, according to a standard operating procedure (SOP) provided by the manufacturer as previously done at the study site when PQ was combined with DP, PA, AL [3], [8], [23].

#### 6.3. Artemether-Lumefantrine (AL)

Participants in the AL arm will be treated with standard doses of AL (Fosun Pharma, Shanghai, China). Tablets containing 20 mg artemether and 120 mg lumefantrine will be administered per manufacturer guidelines (**Table 3**)

**Table 3.** AL dosing

| **Bodyweight (kg)** | **20/120 mg tablet** | | |
| --- | --- | --- | --- |
|  | D0 | D1 | D2 |
| 5 to < 15 kg | 1 disp tab x 2 | 1 disp tab x 2 | 1 disp tab x 2 |
| 15 to < 25 kg | 2 disp tab x 2 | 2 disp tab x 2 | 2 disp tab x 2 |
| 25 to < 35 kg | 3 tab x 2 | 3 tab x 2 | 3 tab x 2 |
| ≥ 35 kg | 4 tab x 2 | 4 tab x 2 | 4 tab x 2 |

#### 6.4. Amodiaquine (AQ)

Participants in the ALAQ and ALAQ-PQ arms will receive AQ as tablets of 153 mg. The weight-based treatment schedule in **table 4** aims for a dosage of approximately 10 mg (7.7-15.3mg)/kg/day, given once or twice daily (together with artemether–lumefantrine) for three days.

**Table 4.** AQ dosing

| **Bodyweight (kg)** | **153 mg tablet** | | | | | |
| --- | --- | --- | --- | --- | --- | --- |
|  | **D0**  **(0hr)** | **D0**  **(8hr)** | **D1 (24hr)** | **D1 (36hr)** | **D2 (48hr)** | **D2 (60hr)** |
| 10 to 19.9 | 1 tab | 0 tab | 1 tab | 0 tab | 1 tab | 0 tab |
| 20 to 29.9 | 1 tab | 1 tab | 1 tab | 1 tab | 1 tab | 1 tab |
| 30 to 54.9 | 2 tab | 1 tab | 2 tab | 1 tab | 2 tab | 1 tab |
| 55 to 80 | 3 tab | 2 tab | 3 tab | 2 tab | 3 tab | 2 tab |

### 7. OUTCOME MEASURES

#### 7.1. Primary outcome measure

Median within person percent change (presented as percent reduction) in mosquito infection rate in infectious individuals from baseline (day 0, pre-treatment) to day 2 post treatment in the ASAQ, ASAQ-PQ, AL, ALAQ and ALAQ-PQ arms.

#### 7.2. Secondary outcome measures

- Median within person percent change (presented as percent reduction) in mosquito infection rate from baseline to all feeding time-points, with comparison within and between arms.
- Median mosquito infection rate at all feeding time-points, with comparison within treatment arms compared to baseline, and between arms.
- Infectivity to mosquitoes at all feeding time-points, with comparison within treatment arms compared to baseline, and between arms.
- Mean oocyst intensity (in all/all infected mosquitoes) at all feeding time-points, with comparison within treatment arms compared to baseline, and between arms.
- Male and female gametocyte prevalence at all time-points, determined by microscopy or molecular assays, with comparison within treatment arms compared to baseline, and between arms.
- Male and female gametocyte density at all time-points, determined by microscopy or molecular assays, with comparison within treatment arms compared to baseline, and between arms.
- Male and female gametocyte sex ratio (proportion male) at all time-points, determined by microscopy or molecular assays, with comparison within treatment arms compared to baseline, and between arms.
- Gametocyte circulation time (cumulative), determined by microscopy or molecular assays, compared between treatment arms.
- Gametocyte area under the curve (cumulative), determined by microscopy or molecular assays, compared between treatment arms.
- Asexual and total parasite prevalence at all time-points, determined by microscopy or molecular assays, with comparison within treatment arms compared to baseline, and between arms.
- Asexual and total parasite density at all time-points, determined by microscopy or molecular assays, with comparison within treatment arms compared to baseline, and between arms.
- Haemoglobin density at all time-points, with comparison within treatment arms compared to baseline, and between arms.
- Median within person percent change (presented as percent reduction) in haemoglobin density from baseline to all time-points, with comparison within and between arms.
- The frequency and prevalence of adverse events (all AE’s, treatment related AE’s, and haematological AE’s) observed up to and including day 2, 7, and 14 post-treatment, and at all timepoints.

#### 7.3. Exploratory outcomes

- Parasite genotype and transcriptional analysis at baseline and at post-treatment timepoints, both for blood stage parasites and oocysts.
- Plasma biomarkers (antibodies and parasite protein) at baseline and at post-treatment timepoints.
- Association between gametocyte density and mosquito infection rates per study drug and time-point, to examine evidence for loss in gametocyte fitness/sterilization.
- Human genotype analysis at baseline (G6PD, CYP2D6, HBB)
- ALT/Creatine density at all time-points with comparison within treatment arms compared to baseline, and between arms.
- Gametocyte density, mosquito infection rate and mean oocyst intensity before and after MACS, for within treatment arm comparison to baseline and comparison between treatment arms.

**Table 5. Outcome measures**

| Outcome measure | Day 0 | Day 1 | Day 2 | Day 7 | Day 14 | Day 21 | Day 28 |
| --- | --- | --- | --- | --- | --- | --- | --- |
| Mosquito infectivity | * |  | * | * | * | ** | ** |
| Asexual parasite prevalence & density | * |  | * | * | * | * | * |
| Gametocyte prevalence, density, sex ratio, genotype | * |  | * | * | * | * | * |
| Biochemistry (ALT/Creatinine) | * |  | * |  | * |  |  |
| CBC | * |  | * |  | * |  |  |
| Haemoglobin | * | * | * | * | * | * | * |
| Safety assessment: signs of haemolysis | * | * | * | * | * | * | * |
| Safety assessment: Adverse Events | * | * | * | * | * | * | * |
| Plasma biomarkers | * |  | * | * | * | * | * |
| Mosquito infectivity after MACS | * |  | * |  |  |  |  |

**** Mosquito infectivity assays will be performed at day 21 for any individuals infectious at day 7 or 14; and at day 28 for any individuals infectious at day 14 or 21.

### 8. PROCEDURES

#### 8.1. Informed consent

Consenting procedures will vary based on age of the potential participant. Participants aged 18 years and above will provide informed consent. For participants under 18 years of age we will seek parental consent. In addition to parental consent, assent will be sought for children aged 10-17 years. The Ethics committee in Mali does not require that participants under the age of 10 provide assent. The written, informed consent procedure will be conducted in French or in a local language understood by subject as described in the information and consent form.

The informed consent document will be used to explain the risks and benefits of study participation to the participant and in the case of participants < 18 years, the participant’s parent in simple terms before the subject is enrolled in the study. The informed consent document contains a statement that the consent is freely given, that the subject is aware of the risks and benefits of entering the study, and that the subject is free to withdraw from the study at any time. Written consent must be given by the participant or the participant’s parent, after the receipt of detailed information on the study. The informed consent form will be signed and personally dated by the participant or the participant’s parent and the person who conducts the informed consent discussion. The original signed informed consent form will be retained in the participant’s chart and another copy will be provided to him. A participant who is unable to read or write will place an imprint of his finger in place of a signature; in addition, an independent witness will sign the consent form to attest that the information in the consent form was orally conveyed to the participant. Specific consent will be provided for sharing samples and anonymized data to collaborating institutes outside Mali and for sharing data in anonymous form on a digital repository.

#### 8.2. Study procedures

Participants who agree to participate and provide written, informed consent will be assessed for the presence of sexual blood stage parasite with a thick blood film. Individuals with ≥16 gametocytes/µL will go through additional screening for eligibility including haemoglobin measurement, CBC, assessment of the renal and liver functions and pregnancy tests. Recruitment will continue until 100 infected individuals with microscopically detectable *P. falciparum* gametocytes (≥16/µL) are recruited and randomised. Individuals with symptomatic malaria will not be enrolled in the study and will be treated with artemether + lumefantrine, the standard treatment for uncomplicated malaria as per the Ministry of Health policy in Mali. Individuals screened found with other acute diseases will also be treated according to the standard of care in Mali. Those with chronic disease will be referred to the district hospital for further evaluation and management.

Eligible patients will then be randomized (see below under randomization procedure) to one of the treatment groups, and drugs will be administered. In case of vomiting within 30 minutes after drug intake, the drugs will be re-administered once.

The participants will be compensated for any travel costs and for work loss income. Participants will be followed for 28 days as described in **Table 6** (below). Single venous draws or finger pricks will be used.

Any individuals determined to have an adverse event (haemolytic or otherwise) will be treated in accordance with national guidelines and best practice and will be withdrawn from the study at the discretion of the study clinician.

**Table 6. Sampling framework for study participants**

| Required vacutainer | Time of sampling  (d = day) | Required volume (mL) | Day 0 | Day 1 | Day 2 | Day 7 | Day 14 | Day 21 | Day 28 |
| --- | --- | --- | --- | --- | --- | --- | --- | --- | --- |
|  | Type of sampling^$^ |  | V | C | V | V | V | V | V |
| Heparin (2-5mL) | Mosquito infectivity | 2 | * |  | * | * | * | ** | ** |
|  | MACS | 3 | * |  | * |  |  |  |  |
| EDTA (0.05-3.4mL) | Blood in RNA protect for RNA analysis/parasite genotyping | 0.7 | * |  | * | * | * | * | * |
|  | Blood smear for asexual parasite and gametocyte density | 0.1 | * |  | * | * | * | * | * |
|  | Filter paper blood spots | 0.05 | * |  | * | * | * | * | * |
|  | Haemoglobin | 0.05 | * | * | * | * | * | * | * |
|  | Plasma | 1 | * |  | * | * | * | * | * |
|  | CBC | 0.5 | * |  | * | * | * |  |  |
|  | Human genotyping | 1 | * |  |  |  |  |  |  |
| SST (0.8 mL) | Biochemistry | 0.8 | * |  | * | * | * |  |  |
|  | **Maximum total Volume of blood sampled (mL)** | **38.65** | **9.2** | **0.05** | **9.2** | **6.2** | **6.2** | **3.9** | **3.9** |

*$ V denotes venous blood sampling and C denotes capillary sampling from a finger prick.*

*** Denotes volume taken for any individuals infectious at day 7 or 14 (day 21) and for any individuals infectious at day 14 or 21 (day 28) .*

#### 8.3. Randomization procedure

Eligible participants will be assigned the next sequentially numbered study ID number based on a pre-printed Study ID List created by a study investigator. Randomization will be computer generated using a 1:1:1:1:1 ratio. The randomisation codes will be provided in opaque sealed envelope to the study pharmacist. The study pharmacist in Mali will open the corresponding sealed, opaque envelope and provide the intervention to study participants. Randomization will be performed in blocks of varying sizes.

#### 8.4. Blinding

This is a single-blind randomised controlled trial. The treating physician and staff involved with assessing all laboratory outcomes of the study are blinded, but no placebo will be used. The study pharmacist will be unblinded and responsible for randomisation and treatment administration. Entomology staff involved in the mosquito feeding assays will be blinded for the parasitology results.

### 9. SAFETY EVALUATION

The major safety endpoint is haemolysis. For this reason, haemoglobin will be measured before treatment, at days 1, 2, 7, 14, 21, and 28. Previous studies have shown that highest haemoglobin fall related to drug induced haemolysis can be detected within 7 days after treatment. In addition at 24 hours, and subsequent days of follow up, a questionnaire assessing adverse events (AE) will be carried out. All participants will have access to contact study medical staff 24 hours a day and medical facilities that can give safe blood transfusions. Participants who require blood transfusions will be hospitalised at the district hospital in Ouelessebougou or referred to the University hospitals in Bamako for proper management. Additional safety precautions due to the small risk of transient haemolysis in this population will include the performance of clinical examinations and monitoring for adverse events (AE) at all sampling time points, carefree of charge to all participants throughout the duration of follow-up.

#### 9.1. Definitions

#### 9.1.1. Adverse events

Adverse events are defined as any undesirable experience occurring to a subject during the study, whether or not considered related to trial. An AE can therefore be any unfavourable and unintended sign (including an abnormal laboratory finding), symptom, or disease (new or exacerbated) temporally associated with a study intervention. AEs may include events that occur as a result of protocol-mandated procedures (i.e. invasive procedures). All AEs reported spontaneously by the subject or observed by the trial clinicians, or their staff will be recorded. Abnormal laboratory findings or other abnormal assessments that are judged by the trial clinicians to be clinically significant will be recorded as AEs or serious adverse events (SAEs) if they meet the definition. The investigators and trial clinicians will exercise their medical and scientific judgement in deciding whether an abnormal laboratory finding, or other abnormal assessment is clinically significant.

#### 9.1.2. Serious adverse events

A serious adverse event is any untoward medical occurrence or effect that at any dose:

- Results in death
- Is life threatening (at the time of the event)
- Requires hospitalisation
- Results in persistent or significant disability or incapacity
- Congenital anomaly/birth defect
- Medically important event

All SAEs will be reported by the principal investigators to the Ethics Committees and DSMB within 48 hours of awareness of the PI.

#### 9.1.3. Unexpected adverse events

An AE is considered unexpected if it is not listed in the Investigator’s Brochure (IB) or Package Insert (for marketed products) or is not listed at the specificity or severity that has been observed.

#### 9.1.4. Serious and Unexpected Suspected Adverse Reaction (SUSAR)

A SUSAR is any AE that is assessed as being serious, is suspected of having a causal relationship to the trial treatment and is unexpected (as per 3.2.4), will require expedited reporting to the Sponsor, RA and IRB/IEC which have approved the clinical trial and all other organisations as required under the terms of the individual contracts (e.g. relevant pharmaceutical companies, NHS Trusts etc.).

Fatal or life-threatening SUSARs must be reported within 7 days of receipt of the report and all other SUSARs within 15 days (see Appendix 3 for reporting procedure for SUSARs for LSHTM sponsored trials).

#### 9.1.5. Serious Unanticipated Problem (UP)

An UP is any event, incident, experience, or outcome that is

1. unexpected in terms of nature, severity, or frequency in relation to
   a. the research risks that are described in the approved research protocol and informed consent document, IB, or other study documents; and
   b. the characteristics of the subject population being studied; and
2. possibly, probably, or definitely related to participation in the research; and
3. places subjects or others at a greater risk of harm (including physical, psychological, economic, or social harm) than was previously known or recognized.

#### 9.1.6. Unanticipated Problem that is not an Adverse Event (UPnonAE)

An UPnonAE is an UP that does not fit the definition of an adverse event, but which may, in the opinion of the investigator, involve risk to the subject, affect others in the research study, or significantly impact the integrity of research data. Such events would be considered a non-serious UP. For example, we will report occurrences of breaches of confidentiality, accidental destruction of study records, or unaccounted-for study drug.

All UPs that are also adverse events will be reported to the sponsor no later than 7 calendar days of site awareness of the event. UPs that are not AEs will also be reported to the sponsor.

#### 9.1.7. Confirmatory pregnancy test at the end of follow-up

Urine and/or serum pregnancy test (β-hCG) will be repeated on the final day of follow-up (day 28) for participants who received a pregnancy test at enrolment. The results will be reported in the communication to the DSMB.

#### 9.2. Adverse Event Data Collection

Safety assessments will be performed and recorded by the trial clinicians. All AEs/reactions, observed by the trial clinicians or by the subject, will be accurately documented in the case report form. For each event/reaction the following details will be recorded:

1.	Description of the event(s)/reaction(s)

2.	Date and time of occurrence

3.	Duration

4.	Intensity

5.	Relationship with the intervention

6.	Action taken, including treatment

7.	Outcome

In addition, symptoms will be ranked as (1) mild, (2) moderate, or (3) severe, depending on their intensity. All AEs except fever will be judged for their intensity according to the following scale:

- Mild (grade 1): awareness of symptoms that are easily tolerated and do not interfere with usual daily activity
- Moderate (grade 2): discomfort that interferes with or limits usual daily activity
- Severe (grade 3): disabling, with subsequent inability to perform usual daily activity, resulting in absence or required bed rest

For fever, the following scale will be used:

•	Mild (grade 1): 37.5 - 38.0°C

•	Moderate (grade 2): > 38.0 to 39.0°C

•	Severe (grade 3): > 39.0°C

When an AE/SAE occurs, it is the responsibility of the principal investigators and trial clinicians to review all documentation (e.g., hospital progress notes, laboratory, and diagnostics reports) related to the event. The principal investigators and trial clinicians will then record all relevant information regarding an AE/SAE on the Case Report Form (CRF) or SAE Report Form, respectively. Furthermore, the trial clinicians will attempt to establish a diagnosis of the event based on signs, symptoms, and/or other clinical information. In such cases, the diagnosis should be documented as the AE/SAE and not the individual signs/symptoms.

#### 9.3. Haematological severe AE

A haematological severe AE is defined as a drop in Hb density of >40% from baseline. These will be reported separately in the data safety and monitoring reports and in any resulting publications, due to their potential linkage with PQ treatment.

#### 9.4. Assessment of causality

The principal investigators and trial clinicians are obligated to assess the relationship between study procedures and the occurrence of each AE/SAE. The clinicians will use clinical judgment to determine the relationship. Alternative causes, such as natural history of the underlying diseases, concomitant therapy, other risk factors and the temporal relationship of the event to the study procedures will be considered and investigated. The relationship of the AE with the study procedures will be assessed considering the factors listed under the following categories:

**Table 7. Causality**

| **Definitely Related** | - reasonable temporal relationship - follows a known response pattern - clear evidence to suggest a causal relationship - there is no alternative aetiology |
| --- | --- |
| **Probably Related** | - reasonable temporal relationship - follows a suspected response pattern (based on similar agents) - no evidence of a more likely alternative aetiology |
| **Possibly Related** | - reasonable temporal relationship - little evidence for a more likely alternative aetiology |
| **Unlikely Related** | - does not have a reasonable temporal relationship   OR   - good evidence for a more likely alternative aetiology |
| **Not Related** | - does not have a temporal relationship   OR   - definitely due to an alternative aetiology |

*Note: Other factors will also be considered for each causality category when appropriate. Causality assessment is based on available information at the time of the assessment of the AE. The investigator may revise the causality assessment as additional information becomes available.*

#### 9.5. Assessment of outcome

Assessment of each AE must be assessed according to the following classification:

**Table 8. Outcome**

| **Completely recovered** | The subject has fully recovered with no observable residual effects |
| --- | --- |
| **Not yet completely recovered** | The subject condition has improved, but still has some residual effects |
| **Deterioration** | The subject’s overall condition has worsened |
| **Permanent damage** | The AE has resulted in a permanent impairment |
| **Death** | The subject died due to the AE |
| **Ongoing** | The AE remains the same as at onset |
| **Unknown** | The outcome of the AE is not known because of lost to follow-up |

All adverse events will be recorded on individual CRF with the following information: the severity grade (mild, moderate, severe), its relationship to the study drug(s) (related/not related), its duration (start and end dates or if continuing at final exam), actions taken, outcome, whether it constitutes a serious adverse event (SAE).

#### 9.6. Data and Safety Monitoring Committee (DSMB)

A Data and Safety Monitoring Committee (DSMB) will be established and convened before the onset of the trial; an independent local safety monitor (LSM) will be appointed in Mali and form part of the DSMB. The DSMB will convene meetings in person or by telephone conference prior to the study initiation, after the first 50 subjects have been enrolled and upon completion of the study. In addition to these three scheduled meetings, *ad hoc* meetings may be organised based on monthly progress reports that are provided to the DSMB. These monthly reports include an update on the number of participants who were enrolled, who completed the study and who discontinued the study (with reason provided). In addition, key safety data are provided: AEs and up to day 14 haemoglobin concentrations. Any SAEs or Suspected Unexpected Serious Adverse Reactions (SUSARs) are first reported and discussed among the research team and LSM and subsequently reported within 48 hours to the complete DSMB. If an SAE or SUSAR is deemed possibly, probably, or definitely related to the study medication by the DSMB, the trial will be stopped until the DSMB has decided how to proceed or to stop the study.

#### 9.7. Treatment failure criteria

In case a patient develops symptomatic malaria (denoted as fever and the presence of asexual parasitaemia denoted by microscopy) at any time during follow-up, they will be treated with a full course of Dihydroartemisinin-Piperaquine. DP treatment tablets contain 40 mg dihydroartemisinin/320 mg piperaquine (Eurartesim, Sigma Tau) and will be administered per manufacturer guidelines shown in **Table 5**.

**Table 9**. DP dosing

| **Body weight (kg)** | **Daily dose (mg)**  **(to be given 1x/day for 3 days)** | | **Tablet strength and number of tablets per dose** |
| --- | --- | --- | --- |
|  | Piperaquine | DHA |  |
| 5 to <7 | 80 | 10 | ½ x 160mg / 20mg tablet |
| 7 to <13 | 160 | 20 | 1 x 160mg / 20mg tablet |
| 13 to <24 | 320 | 40 | 1 x 320mg / 40mg tablet |
| 24 to <36 | 640 | 80 | 2 x 320mg / 40mg tablet |
| 36 to <75 | 960 | 120 | 3 x 320mg / 40mg tablet |
| 75 to 80 | 1,280 | 160 | 4 x 320mg / 40mg tablet |
| >80 | Not eligible | | |

### 10. LABORATORY PROCEDURES

#### 10.1. Blood film

Thick and/or thin blood films for parasite counts will be obtained and examined at screening to confirm *P. falciparum* monoinfection. Giemsa-stained thick and/or thin blood films will be examined at a magnification of 100×. The blood smear will be considered negative if no parasites are seen after examining 100 high powered fields.

#### 10.2. Gametocyte and asexual stage density measurement

Blood slides stained with Giemsa will be double read over 500 fields for quantification of gametocytes and asexual stages. EDTA samples of blood (200 µL) will be tested for molecular quantification of gametocytes and asexual parasites. Parasite DNA and RNA will be extracted from whole blood samples in EDTA tubes and tested using quantitative real-time polymerase chain reaction (qPCR) with a detection limit of 0.02-0.1 parasites/µL of blood and highly precise parasite quantification [24]. Quantification of gametocytes will be based on sex specific female (Pfs25/CCP4) and male (PfMGET) mRNA detection and quantification by reverse transcriptase-PCR [25]. Markers of asexual stage parasites will include 18s Ribosmal and SBP1 mRNA. Markers of sexual commitment will also be assessed (e.g. Ap2-g).

Further molecular analysis of samples from individuals who have undergone feeding assays (either from whole blood samples or from mosquito midguts) will include parasite genotyping (e.g. to assess clonal complexity or determine the presence of specific deletions e.g. HRP2) and human genotyping for specific genes (HBB, CYP2D6, G6PD).

#### 10.3. Serology and biomarkers

Serological analysis of these samples will include assessment of antibody responses to gametocyte and asexual stage proteins in the microarray platform [26]. Non antibody protein biomarker assessment will be conducted using bead-based assays to assess for correlates of infection and infectivity (i.e. proteins associated with inflammation), or parasite density (HRP2), or parasite proteins linked with gamete activation in the mosquito midgut (i.e Pfg377). If plasma remains after mosquito feeding, some may be stored and used in standard membrane feeding assays in Radboud University, Nijmegen, the Netherlands, for the assessment of functional transmission reducing immunity [27].

#### 10.4. Mosquito infectivity assay

For each assessment of infectivity 2 ml of heparinized blood will be drawn from the study participant and stored at 37^o^C and transported to the insectary. At the insectary, using standard procedures, ~70 *A. gambiae* will be fed on the subjects’ blood for 15-20 minutes (this figure is based on a previous study where 61 fed mosquitoes allowed for an average of 50 mosquitoes surviving until day 7 after the feeding experiment and being available for dissection and examination for oocysts). All of these mosquitoes will be dissected on the 7^th^ day after the feeding assay for prevalence of mosquitoes with oocysts and quantification of oocysts. Infected guts will be stored for later PCR confirmation of oocysts.

#### 10.5. Magnetic-activated cell sorting

Blood samples will be enriched for gametocytes using MACS LS columns as previously described [28].

#### 10.6. Haemoglobin concentrations

Haemoglobin concentrations will be also measured regularly throughout the study using the HemoCue system (Hemocue AB, Angelholm, Sweden).

#### 10.7. Complete blood count

Using standard techniques, the clinical laboratory will perform the complete blood count (CBC) and platelet count. The following CBC parameters will be evaluated for exclusion criteria at baseline and for safety assessment during the study on days 2, 7 and 14: white blood cells, haemoglobin, haematocrit, platelets.

#### 10.8. Biochemistry

Blood chemistries values will be also measured throughout the study. The following parameters will be evaluated for exclusion criteria at baseline and for safety assessment during the study on days 2, 7 and 14: serum creatinine and ALT.

### 11. DATA MANAGEMENT AND ANALYSIS

All data will be stored on secure password protected databases. Data that are collected on paper forms will be double entered. Data collected through handheld devices or directly produced by laboratory equipment will be examined for quality assurance and fed into the database using double data entry. Handheld devices used for data input and temporary storage will be password protected and data will be encrypted.

#### 11.1. Analysis plan

Mosquito infectivity will be assessed at three levels: the mean number of oocysts in a sample of mosquitoes (i.e. oocyst intensity), the proportion of mosquitoes infected with any number of oocysts (i.e. mosquito infection rate), and the infectivity of the study participant to any number of mosquitoes (i.e. infectivity to mosquitoes). The primary outcome measure will be the percent change in mosquito infection rate within each arm by day 2 compared to baseline, with other time-points of infectivity assessments as secondary outcomes. Percent change will be reported as percent reduction (with 100% as total reduction of transmission, and negative values as enhanced transmission). Other secondary transmission outcomes are as described in the ‘Objectives’ section above. Some exploratory objectives may be published separately from the main primary/secondary outcomes.

Statistical analysis will be conducted using the most recent STATA and SAS versions (16.0 and SAS version 9.4 at time of writing). Mosquito infection data will be analysed at time-points after baseline only for those individuals who are infectious at baseline, although the data and analyses from all enrolled individuals will be published in the appendix for completeness. Individuals will be classified as infectious to mosquitoes if they infect at least 1 mosquito, with any number of oocysts. Infectivity to mosquitoes and parasite/gametocyte prevalence will be compared within and between treatment arms using generalised linear models (family: binomial, z-score, co-efficient with 95% CI) or fishers exact tests. Absolute haemoglobin density and percent change in haemoglobin density (relative to baseline) will be compared using paired t-tests (t-score for difference compared to day 0) and two-way t-tests (t-score for difference between ACT matched treatment groups at each time-point). The proportion of gametocytes that are male will be calculated for all values with total gametocyte densities of 0.2/µL and over. Gametocyte circulation time will be calculated to determine the average period (in days) that a mature gametocyte circulates in the blood prior to clearance, using a deterministic compartmental model that assumes a constant rate of clearance and has a random effect to account for repeated measures on individuals, as described previously [29]; circulation time will be analysed using t-tests (t score for difference between ACT matched treatment groups) or linear regression analyses. Area under the curve (AUC) of gametocyte density per participant over time will be calculated using the linear trapezoid method [30] using the first 28 days of observation only, and analysed by fitting linear regression models to the log10 adjusted AUC values, with and without adjustment for baseline gametocyte density (t-score, coefficient with 95% CI). All other analyses of quantitative data will be performed using Wilcoxon sign rank tests (z-score for difference compared to matched values at day 0) and Wilcoxon rank-sum tests (z-score for difference between ACT matched treatment arms at each time-point). The nonparametric van Elteren’s test, an extension of the Wilcoxon rank sum test, will be used to compare the infectivity between treatment arms after adjusting for gametocyte density, as previously described [19]. This will compare infectivity between groups after stratifying into categories based on total gametocytes per microliter. Within these bins differences in infectivity between arms will be tested, thereby investigating sterilizing effects of gametocytocidal drugs. For all analyses, the threshold for statistical significance will be set at p<0.05.

#### 11.2. Publication/Dissemination

Findings will be analysed and presented to local researchers and stakeholders in Mali and at other research meetings and symposium. Data will be submitted for publication in peer reviewed journals in due course. Anonymized data will be shared in public repositories upon publication.

### 12. ETHICAL CONSIDERATIONS

This research will be conducted in compliance with the protocol, Good Clinical Practice (GCP) guidelines, and all applicable regulatory requirements. A copy of the protocol, informed consent forms, and any other documents given to study participants will be submitted to the ethics committees (EC) of all the institutions involved. Written approval will be obtained for all subsequent amendments to the protocol, informed consent documents, and other study documentation referenced above. The investigator will notify the ECs and DSMB of violations of the protocol and serious adverse events. Ethical approval will be sought from the Ethics Committee of the Faculty of Medicine, Pharmacy, and Dentistry of the University of Science, Techniques, and Technologies of Bamako (Bamako, Mali) and the Research Ethics Committee of the London School of Hygiene & Tropical Medicine (London, UK). No activities will start before approval has been received by both committees.

#### 12.1. Confidentiality

The investigator will ensure that the subject’s anonymity is maintained. Participants will not be identified in any publicly released reports of this study. All records will be kept confidential to the extent provided by laws and regulations. The study monitors and other authorized representatives of the regulatory authorities may inspect all documents and records required to be maintained by the Investigator.

All laboratory specimens, evaluation forms, reports, and other records that leave the site will be identified only by a coded number to maintain subject confidentiality.

Fully anonymized data will be uploaded to publicly available data repositories (e.g. https://datadryad.org/, https://www.biorxiv.org/) if requested as a pre-requisite for publication of this research. This will be explained in the participant information and during the consenting procedure. The upload of anonymized data will only be permitted if the participants/participants parents provide explicit approval during consent (a tick box in addition to the signature required for trial participation).

#### 12.2. Risk

This study only involves the use of antimalarial drugs at doses equal or lower than the recommended dosing and schedules. All drugs have been studied thoroughly and their toxicities are well described.

Venipuncture can be associated with pain and bruising at the site of the prick, and rarely infection. Risks will be minimized by the study sites by ensuring adequate training of staff in all procedures and supply of consumables. The study site adheres to the standards of GCP and will be monitored.

#### 12.2.1. Risk of artesunate-amodiaquine

About 30% of treated patients experience adverse reactions. Most of the reported adverse reactions are similar to symptoms usually seen during a malaria attack. Common side effects of artesunate-amodiaquine include anorexia, abdominal pain, nausea, asthenia, somnolence, insomnia and cough. More serious adverse reactions, although infrequently, are anaemia and vertigo.

#### 12.2.2. Risk of primaquine

Primaquine is known to cause dose-dependent haemolysis among individuals who are deficient in glucose-6-phosphate dehydrogenase (G6PD), an enzyme involved in glucose metabolism prevalent in malaria endemic geographies [31]. Haemolysis in G6PD-deficient individuals is caused by oxidative stress induced by primaquine within red blood cells (RBCs); the extent of haemolysis depends on several factors including the dose of primaquine, the type of G6PD deficiency, the functional G6PD assessment at the time of dosing, the concurrent use of other oxidant drugs, and whether the person is hemizygous (homozygous) or heterozygous for G6PD-deficiency [31]. In 2010 the WHO recommended the addition of a single low dose of primaquine (0.25 mg/kg) to standard ACT treatment, without mandatory G6PD testing to block *P. falciparum* transmission in areas threatened by artemisinin resistance or approaching malaria elimination [1], [32]. The safety of single low dose PQ has subsequently been confirmed in clinical trials where PQ was combined with either DP or AL [33]–[35]; two of these studies were specifically designed to assess safety in G6PD deficient individuals and found no clinically relevant reductions in haemoglobin concentrations [33], [34]. PQ use is now widespread throughout the world, and many countries have adopted the 0.25 mg/kg WHO recommendation for single low dose PQ without prior G6PD testing [36]. Common side effects include heartburn, nausea and abdominal cramps.

#### 12.2.3. Risk of artemether-lumefantrine

Reported AL side effects have generally been mild. The main side effects are GI upset: anorexia (~18%), nausea (~5%), vomiting (~18%), abdominal pain (~5%), and diarrhoea (~10%), headache (~10%), dizziness (~4%), fatigue (~1%) and sleep disturbance (~2%). Other symptoms reported infrequently include palpitations, myalgia, arthralgia (all of which could be disease related), and rash.

#### 12.2.4. Risk of amodiaquine

The main side-effects of amodiaquine are nausea, vomiting and fatigue which are mild to moderate in nature. When the drug was used for prophylaxis, rare adverse reactions of agranulocytosis and hepatotoxicity were observed.

#### 12.2.5. Risk of artemether-lumefantrine-amodiaquine

In two recent studies [5], [6], artemether-lumefantrine-amodiaquine was found to be generally well tolerated and safe, although mild adverse events (mainly gastrointestinal) were more frequent compared to artemether-lumefantrine alone. The addition of amodiaquine was also observed to cause a small prolongation of the electrocardiogram corrected QT interval compared to artemether-lumefantrine alone (mean increase at 52 h of 8.8 ms versus 0.9 ms), but not to the extent of cardiac arrhythmias. In addition, amodiaquine use was associated with a bigger decrease in heart rate compared to artemether-lumefantrine alone and a higher incidence of bradycardia (<54 beats/minute), both in adults and children.

#### 12.3. Benefits

The benefits of the participation include free treatment of malaria and other acute diseases that may occur during the follow-up period. Participants who are in the groups that receive primaquine may be less infectious to mosquitoes and reduce the chance of having mosquitoes in their environment becoming infectious. Findings will help in the treatment of future patients and communities with malaria and further optimize the design of community intervention programmes.

#### 12.4. Compensation

Participants will receive compensation for the time and travel for protocol specified visits. This compensation will be carefully evaluated and provided upon agreement of the local ethics committee. In Mali, the estimated cost of time and travel expense per visit in Ouelessebougou for residents of Ouelessebougou is estimated to be 1500 CFA (~3 US $). The compensation for enrolment visit will be 3000 CFA (~6.0 US $) and 1500 CFA (~3 US $) for each additional visit.

### 13. USE AND STORAGE OF STUDY SAMPLES

Samples collected will be stored at MRTC in Bamako, Mali and some will also be shipped to collaborating research centres such as the London School of Hygiene & Tropical Medicine and/or Radboud University medical centre to perform specific tests as described above. Samples may be kept for a maximum of 10 years. As described in the patient consent forms secondary use of patient samples may be shared with collaborating institutes, if ethical approval and appropriate material transfer agreements are in place.

### 14. SPONSOR INFORMATION

#### 14.1. Indemnity

London School of Hygiene & Tropical Medicine holds Public Liability ("negligent harm") and Clinical Trial ("non-negligent harm") insurance policies which apply to this trial.

#### 14.2. Sponsor

London School of Hygiene & Tropical Medicine will act as the main sponsor for this study. Delegated responsibilities will be assigned locally.

#### 14.3. Audits and Inspections

The study may be subject audit by the London School of Hygiene & Tropical Medicine under their remit as sponsor, the Study Coordination Centre, and other regulatory bodies to ensure adherence to GCP.

### 15. REFERENCES

[1]	World Health Organization, ‘Updated WHO policy recommendation October 2012: single dose primaquine as a gametocytocide in Plasmodium falciparum malaria’. 2012.
